# Schizophrenia-associated polygenic liability and structural genomic risk demonstrate broadly distributed neuropsychiatric associations in the *All of Us* Research Program

**DOI:** 10.64898/2026.09.21.26363600

**Authors:** Anne Marie Wells, Annie Dang, Agustin Ruiz

## Abstract

**Background:** Schizophrenia liability reflects both the aggregate effects of many common genetic variants and rare structural variation at recurrent neurodevelopmental copy-number variant (ND-CNV) loci. Although both components of genetic liability have been associated with schizophrenia and related phenotypes, it remains unclear whether they exhibit similar neuropsychiatric association profiles or show evidence of interaction when examined jointly.

**Methods:** We analyzed 71,992 participants from the *All of Us* Research Program Controlled Tier version 8 dataset, including 2,115 participants carrying structural variants overlapping curated recurrent ND-CNV loci. We integrated binary ND-CNV carrier status, an ancestry-adjusted and standardized reduced schizophrenia polygenic risk score (PRS), and longitudinal electronic health record-derived psychiatric and neurodevelopmental phenotypes. Logistic regression models evaluated pooled, deletion-overlapping, and duplication-overlapping carrier associations, schizophrenia PRS effects, and PRS × ND-CNV interactions. Dimensional analyses, confirmatory factor analysis (CFA), and structural equation modeling (SEM) provided complementary summaries of phenotype covariance and assessed the robustness of primary associations across alternative representations of the phenotype space.

**Results:** Schizophrenia PRS and ND-CNV carrier status showed distributed association profiles extending beyond schizophrenia diagnosis across psychosis-spectrum, neurodevelopmental, cognitive, and internalizing phenotypes. ND-CNV carriers had higher estimated odds of schizophrenia diagnosis than non-carriers (OR = 1.49, 95% CI 1.09–2.03), while each 1-SD increase in schizophrenia PRS was associated with higher odds of a broad psychosis-spectrum phenotype (OR per SD = 1.07, 95% CI 1.03–1.10). Deletion-overlapping carriers exhibited generally larger and more consistently positive association estimates than duplication-overlapping carriers. Psychosis-spectrum prevalence increased from 5.4% in the lowest PRS decile to 6.7% in the highest, while ND-CNV carriers maintained consistently higher prevalence than non-carriers across the PRS distribution. PRS × ND-CNV interaction estimates remained near the null, providing little evidence of widespread multiplicative interaction. Among tested covariance structures, bifactor models showed the lowest residual misfit and were therefore carried forward for SEM analyses.

**Conclusions:** Schizophrenia-associated polygenic liability and structural variation overlapping recurrent ND-CNV loci exhibit distributed neuropsychiatric association profiles with limited evidence of widespread multiplicative interaction. Their joint evaluation in the large, ancestrally diverse *All of Us* cohort demonstrates that distinct forms of genomic liability can be examined within a shared, real-world phenotype space extending beyond schizophrenia diagnosis alone.

## INTRODUCTION

Schizophrenia is clinically defined primarily by psychotic symptoms, yet its phenotypic expression extends well beyond psychosis to cognitive dysfunction, developmental differences, functional impairment, and psychiatric comorbidity^1,2^. Substantial heterogeneity ^2–4^ exists across affected individuals in symptom profiles, cognitive trajectories, developmental histories, psychiatric comorbidities, and treatment response^5,6^. Increasing genetic evidence suggests that this clinical heterogeneity reflects genetic influences extending beyond schizophrenia diagnosis itself^1,7–10^. That genetic variants associated with schizophrenia have been linked to a wide range of neurodevelopmental, cognitive, and psychiatric traits raises a fundamental question: how is schizophrenia-associated genetic risk expressed across the neuropsychiatric phenotype space, and do distinct forms of inherited genetic liability produce similar or divergent patterns of clinical association?

Insights into these questions have emerged through the study of both common and structural forms of schizophrenia-associated genetic susceptibility. Schizophrenia, like many psychiatric disorders, is highly polygenic^7,11–15^, with risk distributed across numerous common variants whose aggregate effects can be quantified using polygenic risk scores (PRS). Some of the strongest known genetic risk factors for schizophrenia are recurrent neurodevelopmental copy-number variants (ND-CNVs)^16–32^, which are large structural variants occurring at recurrent genomic loci that have been repeatedly associated with disorders of neurodevelopment in clinically ascertained cohorts. Schizophrenia PRS and recurrent ND-CNVs occupy contrasting ends of the schizophrenia-associated genetic risk spectrum: PRS captures the cumulative effects of common variants of individually small effect, whereas carrying a large deletions and duplications overlapping recurrent ND-CNV loci is rare in the population but can confer substantially larger individual effects^8,14,33,34^. Despite these differences, both have been associated with overlapping psychiatric, cognitive, and neurodevelopmental phenotypes^22,34–36^, suggesting that schizophrenia-associated genetic risk extends beyond schizophrenia diagnosis itself^37–41^. This convergence raises the possibility that these distinct forms of genetic risk influence partially shared biological systems. If so, common and rare schizophrenia-associated genetic risk should exhibit structured, distributed patterns of association across related neuropsychiatric phenotypes rather than effects narrowly confined to psychotic-spectrum illness.

The coexistence of these complementary forms of genetic liability for schizophrenia provides a unique opportunity to determine whether fundamentally different genetic architectures produce shared or distinct patterns of neuropsychiatric disease. Recent studies^14,41–43^ integrating PRS with clinical and population-based cohorts demonstrated that schizophrenia-associated genetic risk is distributed across multiple psychiatric outcomes rather than confined to schizophrenia diagnosis alone. Similarly, structural variants at recurrent ND-CNV loci have been associated with broad neuropsychiatric phenotypes extending beyond psychosis, ^17,20,21,23,28,29,44–46^. However, relatively few studies^42,47,48^ have jointly evaluated common variants and ND-CNVs for schizophrenia-associated genetic risk within the same population while examining whether their effects converge, diverge, or interact across a wide spectrum of psychiatric outcomes. Because psychiatric disorders frequently co-occur and exhibit substantial phenotypic and genetic overlap, covariance modeling provides a complementary framework for summarizing these broader patterns of phenotype organization^14,49–53^.

In the present study, we integrated recurrent ND-CNV carrier status, a computationally tractable reduced implementation of schizophrenia PRS, and longitudinal electronic health record (EHR)-derived neuropsychiatric phenotypes within the ancestrally diverse *All of Us* Research Program^54^ to characterize how schizophrenia-associated common genetic risk and recurrent ND-CNV loci are distributed across psychiatric and neurodevelopmental outcomes. To complement regression analyses, we incorporated dimensional and covariance modeling to summarize broader patterns of psychiatric phenotype covariance. We hypothesized that schizophrenia polygenic risk and recurrent ND-CNV carrier status would contribute largely to independent associations across a diverse population cohort.

## RESULTS

### Curated recurrent ND-CNV framework overlaps established schizophrenia and neurodevelopmental genomic resources

We first benchmarked the curated recurrent neurodevelopmental copy-number variant (ND-CNV) framework by quantifying interval- and participant-level overlap with established schizophrenia and neurodevelopmental genomic resources before examining phenotype associations (**Figure 1**). The integrated analytic cohort comprised 71,992 participants, including 2,115 recurrent ND-CNV carriers, of whom 1,005 carried deletion-overlapping variants and 1,786 carried duplication-overlapping variants. Because deletion-overlapping and duplication-overlapping classifications were not mutually exclusive, participants carrying both event classes contributed to both direction-specific groups. The curated framework comprised 96 direction-specific deletion and duplication locus definitions corresponding to 51 unique autosomal genomic intervals distributed across the autosomes, including genomic intervals repeatedly implicated in schizophrenia, autism spectrum disorder, intellectual disability, developmental delay, epilepsy, and related neuropsychiatric disorders ^17,23,28,29,44,55–57^ (**Figure 1A**).

**Figure 1.**
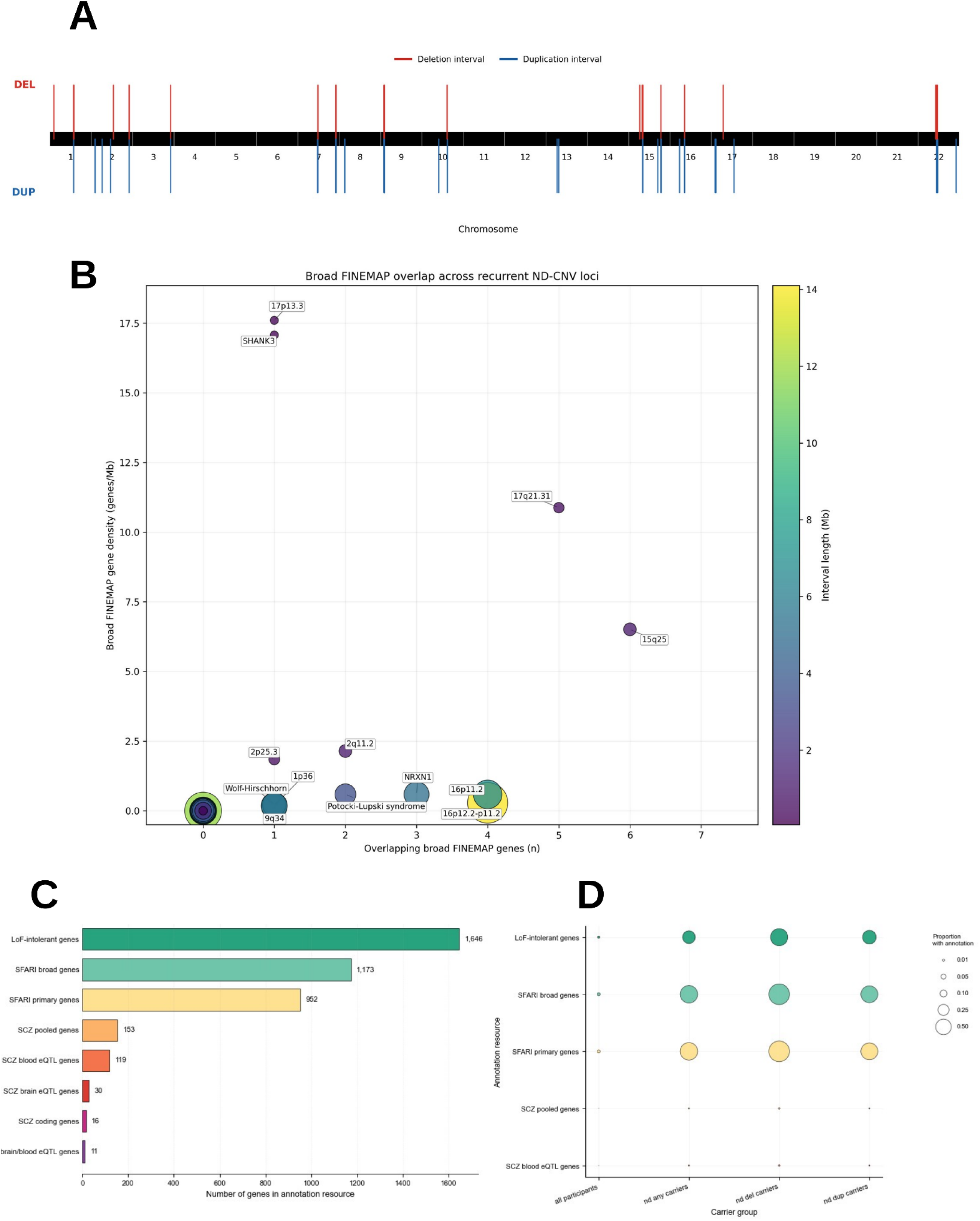
Curated recurrent neurodevelopmental copy-number variant framework and overlap with schizophrenia and neurodevelopmental genomic annotation resources. **(A)** Chromosomal representation of the curated recurrent neurodevelopmental copy-number variant (ND-CNV) framework used for downstream analyses. Black interval bars denote harmonized recurrent deletion and duplication loci curated from established neurodevelopmental and psychiatric genetics literature and converted to GRCh38 genomic coordinates. Primary downstream analyses assigned participant carrier status using reciprocal overlap thresholds ≥30% (ro30). **(B)** Locus-level overlap between curated recurrent ND-CNV intervals and the broad schizophrenia GWAS discovery 435-gene set derived from FINEMAP fine-mapping analyses reported by Trubetskoy *et al.* Each bubble represents one unique autosomal recurrent ND-CNV interval. The x-axis denotes the number of overlapping broad schizophrenia-associated FINEMAP genes within each interval, the y-axis denotes FINEMAP gene density normalized by interval length (genes/Mb), and both bubble size and color represent interval length (Mb). Labels identify recurrent ND-CNV intervals containing one or more overlapping broad FINEMAP genes. **(C)** Gene-level overlap between curated recurrent ND-CNV intervals and 153 high-confidence prioritized gene set derived from schizophrenia and neurodevelopmental genomic annotation resources. Bars summarize the total number of genes intersecting curated ND-CNV intervals across loss-of-function intolerant genes derived from the Genome Aggregation Database (gnomAD) LOEUF framework, Simons Foundation Autism Research Initiative (SFARI) primary autism-associated genes, broader SFARI neurodevelopmental genes, and schizophrenia-prioritized genes aggregated from coding and expression quantitative trait locus (eQTL)-linked resources. **(D)** Participant-level overlap between recurrent ND-CNV carrier intervals and schizophrenia and neurodevelopmental genomic annotation resources. Bars represent the proportion of recurrent ND-CNV carriers whose intervals overlapped at least one gene within each annotation resource. Separate bars are shown for pooled recurrent ND-CNV carriers, deletion-overlapping carriers, and duplication-overlapping carriers. Annotation categories with zero overlap were omitted for visualization clarity.

We next evaluated locus-level overlap between the curated recurrent ND-CNV intervals and the broader schizophrenia GWAS landscape using the FINEMAP-derived discovery gene set reported by Trubetskoy et al.^1,7^ Of 435 broad schizophrenia-associated protein-coding genes, 429 were successfully mapped to the GRCh38 reference annotation, of which 28 (6.5%) intersected at least one curated recurrent ND-CNV interval (**Figure 1B**). Conversely, 13 of 51 unique autosomal recurrent ND-CNV intervals (25.5%) contained at least one broad schizophrenia-associated FINEMAP gene.

The number of overlapping FINEMAP genes varied substantially across recurrent loci and generally increased with interval size, while normalization by interval length demonstrated marked heterogeneity in schizophrenia-associated gene density across recurrent ND-CNV regions. We next examined overlap between the curated intervals and established schizophrenia and neurodevelopmental genomic resources. The 96 recurrent ND-CNV loci intersected 1,646 loss-of-function intolerant genes^58^, 952 SFARI^59^ primary autism-associated genes, 1,173 broader SFARI neurodevelopmental genes, and 153 schizophrenia-prioritized genes aggregated across coding and eQTL-linked resources^1,7^ (**Figure 1C**).

Participant-level analyses demonstrated frequent overlap between participant-carried recurrent ND-CNV intervals and established schizophrenia and neurodevelopmental genomic resources (**Figure 1D**). Among 2,115 recurrent ND-CNV carriers, participant-carried intervals overlapped at least one SFARI primary autism-associated gene in 1,329 individuals (62.8%), a loss-of-function intolerant gene in 695 (32.9%), and at least one schizophrenia-relevant or neurodevelopmental annotation resource in 1,403 (66.3%). Deletion-overlapping carrier intervals more frequently overlapped these annotation resources than duplication-overlapping carrier intervals. Specifically, 88.0% of deletion-overlapping carriers overlapped a broad SFARI neurodevelopmental gene and 91.1% overlapped at least one schizophrenia-relevant or neurodevelopmental annotation resource, compared with 58.4% and 61.5%, respectively, among duplication-overlapping carriers.

### Deletion- and duplication-overlapping ND-CNV carriers exhibit distinct neuropsychiatric association profiles

We next examined how recurrent ND-CNV carrier status was distributed across neuropsychiatric phenotypes using pooled and direction-specific carrier models (**Figure 2A**). The full cohort included 21,607 individuals with mood disorders, 21,767 with anxiety disorders, 4,401 with broad psychosis-spectrum phenotypes, 3,310 with post-traumatic stress disorder (PTSD), 2,248 with attention-deficit/hyperactivity disorder (ADHD), 219 with intellectual disability, 155 with autism spectrum disorder, and 120 with developmental delay.

**Figure 2.**
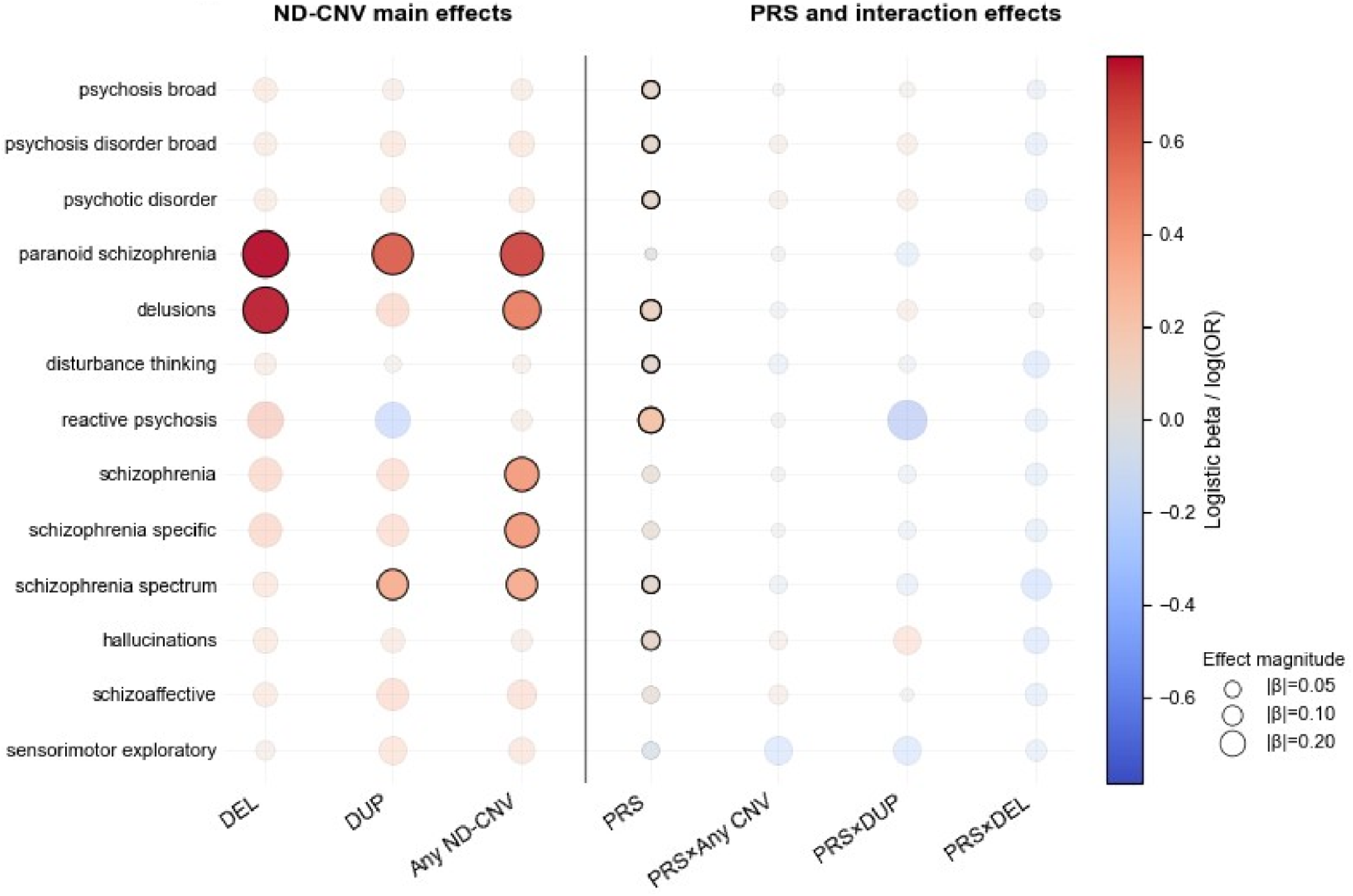
Distributed phenotype-level neuropsychiatric associations across recurrent ND-CNV carriers and schizophrenia polygenic risk. Phenotype-level and dimensional association analyses across 71,992 *All of Us* participants, including 2,115 recurrent ND-CNV carriers. Phenotype-level association profiles across pooled ND-CNV carrier status, deletion-overlapping ND-CNV carrier status (DEL), duplication-overlapping ND-CNV carrier status (DUP), schizophrenia polygenic risk scores (PRS), and PRS × ND-CNV interaction models. Dot size reflects relative standardized effect magnitude. Color denotes association direction (positive or negative), while color intensity reflects standardized effect magnitude. Left panels summarize pooled and direction-specific recurrent ND-CNV association profiles, whereas right panels summarize schizophrenia PRS and PRS × ND-CNV interaction models.

Deletion-overlapping and duplication-overlapping ND-CNV carrier groups exhibited distinct neuropsychiatric association profiles across the modeled phenotype space (**Figure 2A; Figure S1A**). Among pooled recurrent ND-CNV carriers, associations were observed for schizophrenia diagnosis (OR = 1.49, 95% CI 1.09–2.03, *p* = 0.012), schizophrenia-spectrum disorders (OR = 1.38, 95% CI 1.04–1.82, *p* = 0.026), paranoid schizophrenia (OR = 1.97, 95% CI 1.20–3.24, *p* = 0.0078), and delusions (OR = 1.64, 95% CI 1.05–2.56, *p* = 0.028).

Direction-specific analyses showed that deletion-overlapping carrier status generally yielded larger estimated associations than duplication-overlapping carrier status across psychosis-spectrum phenotypes. For example, deletion-overlapping carriers demonstrated a larger estimated association with delusions (OR = 2.13, 95% CI 1.22–3.73, *p* = 0.0079) than duplication-overlapping carriers (OR = 1.47, 95% CI 0.87–2.46, *p* = 0.148), while both carrier groups were associated with paranoid schizophrenia (deletion-overlapping: OR = 2.19, 95% CI 1.11–4.31, *p* = 0.023; duplication-overlapping: OR = 1.83, 95% CI 1.04–3.22, *p* = 0.037). Across the full phenotype panel, deletion-overlapping carrier models also exhibited a greater number of positive association estimates and generally larger effect sizes than duplication-overlapping carrier models (**Figure 2A; Figure S1A**).

### Limited interaction between polygenic and structural genetic risk

We next evaluated whether schizophrenia polygenic risk and recurrent ND-CNV carrier status demonstrated independent or interactive associations across the neuropsychiatric phenotype space (**Figure 2A**). Because schizophrenia PRS was implemented using a computationally reduced scoring framework^15,60^, we first confirmed that the harmonized score preserved the expected genome-wide weight distribution and phenotypic signal (**Figure S2**).

Across phenotype-level analyses, schizophrenia PRS demonstrated broadly distributed positive associations extending beyond schizophrenia diagnosis into multiple psychosis-spectrum phenotypes (**Figure 2A**). Representative associations per 1-SD increase in schizophrenia PRS included broad psychosis-spectrum phenotypes (OR per SD = 1.07, 95% CI 1.03–1.10, *p* = 1.0 × 10⁻⁴), schizophrenia-spectrum disorders (OR per SD = 1.06, 95% CI 1.00–1.12, *p* = 0.032), delusions (OR per SD = 1.12, 95% CI 1.02–1.23, *p* = 0.022), and hallucinations (OR per SD = 1.07, 95% CI 1.00– 1.15, *p* = 0.046). In contrast, recurrent ND-CNV carrier models generally demonstrated larger but less consistently significant pooled associations across the same phenotypes. PRS × ND-CNV interaction terms remained uniformly centered near the null; for broad psychosis-spectrum phenotypes, the ratio of per-SD PRS odds ratios between carriers and non-carriers was 0.996 (95% CI 0.832–1.192, *p* = 0.963).

Psychosis-spectrum prevalence increased progressively across schizophrenia PRS deciles in both recurrent ND-CNV carriers and non-carriers, increasing from 5.4% in the lowest decile to 6.7% in the highest (22.8% relative increase; **Figure S3A**). Mean observed neuropsychiatric phenotype burden similarly increased from 0.73 to 0.77 modeled phenotypes (6.3% relative increase; **Figure S3B**), while the prevalence of elevated neuropsychiatric burden (≥2 modeled phenotypes) increased from 22.7% to 24.2% (6.7% relative increase; **Figure S3C**). Across the PRS distribution, recurrent ND-CNV carriers consistently exhibited higher psychosis prevalence than non-carriers; however, the separation between carrier groups remained relatively stable rather than widening at higher PRS deciles (**Figure S3D**).

Across multiple sensitivity analyses, the primary phenotype-level findings remained robust (**Figure S1B; Figure S4**). Restricting psychosis-spectrum phenotypes to individuals with two or more qualifying diagnostic encounters produced highly similar schizophrenia PRS, recurrent ND-CNV carrier status, and PRS × ND-CNV interaction effect estimates (**Figure S1B**). Likewise, perturbation of demographic, ancestry, healthcare-utilization, and longitudinal ascertainment covariates produced only modest changes in individual effect estimates, with the greatest variation observed following perturbation of ancestry principal components 4 and 5 (**Figure S4**).

### Complementary analytical frameworks support primary genetic association findings

To determine whether the primary phenotype-level associations were preserved after aggregation into broader neuropsychiatric domains, we next summarized the analyses using Research Domain Criteria (RDoC)-informed dimensions (**Figure S5**). Associations per 1-SD increase in schizophrenia PRS demonstrated positive associations across multiple domains, with the largest positive estimates observed for negative valence systems (OR per SD = 1.03, 95% CI 1.01–1.04, *p* = 0.002), cognitive systems (OR per SD = 1.03, 95% CI 1.01–1.05, *p* = 0.004), and positive valence systems (OR per SD = 1.03, 95% CI 1.01–1.05, *p* = 0.014). Pooled recurrent ND-CNV carrier status demonstrated comparatively modest associations across the six RDoC domains, none of which individually reached statistical significance (all *p* > 0.18). However, direction-specific radar plots recapitulated the phenotype-level findings, with deletion-overlapping carrier models exhibiting broader and more uniformly positive associations than duplication-overlapping carrier models across cognitive, negative valence, and positive valence domains (**Figure S5C–D**).

To further evaluate whether the primary genetic association patterns were preserved under alternative representations of the phenotype space, we next performed confirmatory factor analysis (CFA) and structural equation modeling (SEM) (**Figure 3**). Comparison of alternative CFA structures suggested progressive improvement in model fit from one-factor (CFI = 0.996, TLI = 0.994, RMSEA = 0.017 [95% CI 0.016–0.018], SRMR = 0.125) to correlated two-factor (CFI = 0.998, TLI = 0.996, RMSEA = 0.013 [95% CI 0.012–0.015], SRMR = 0.092) to bifactor models (CFI = 0.999, TLI = 0.998, RMSEA = 0.010 [95% CI 0.008–0.012], SRMR = 0.065) (**Figure S6A–B**). Bifactor models exhibited the lowest residual misfit and were used for subsequent SEM analyses.

**Figure 3.**
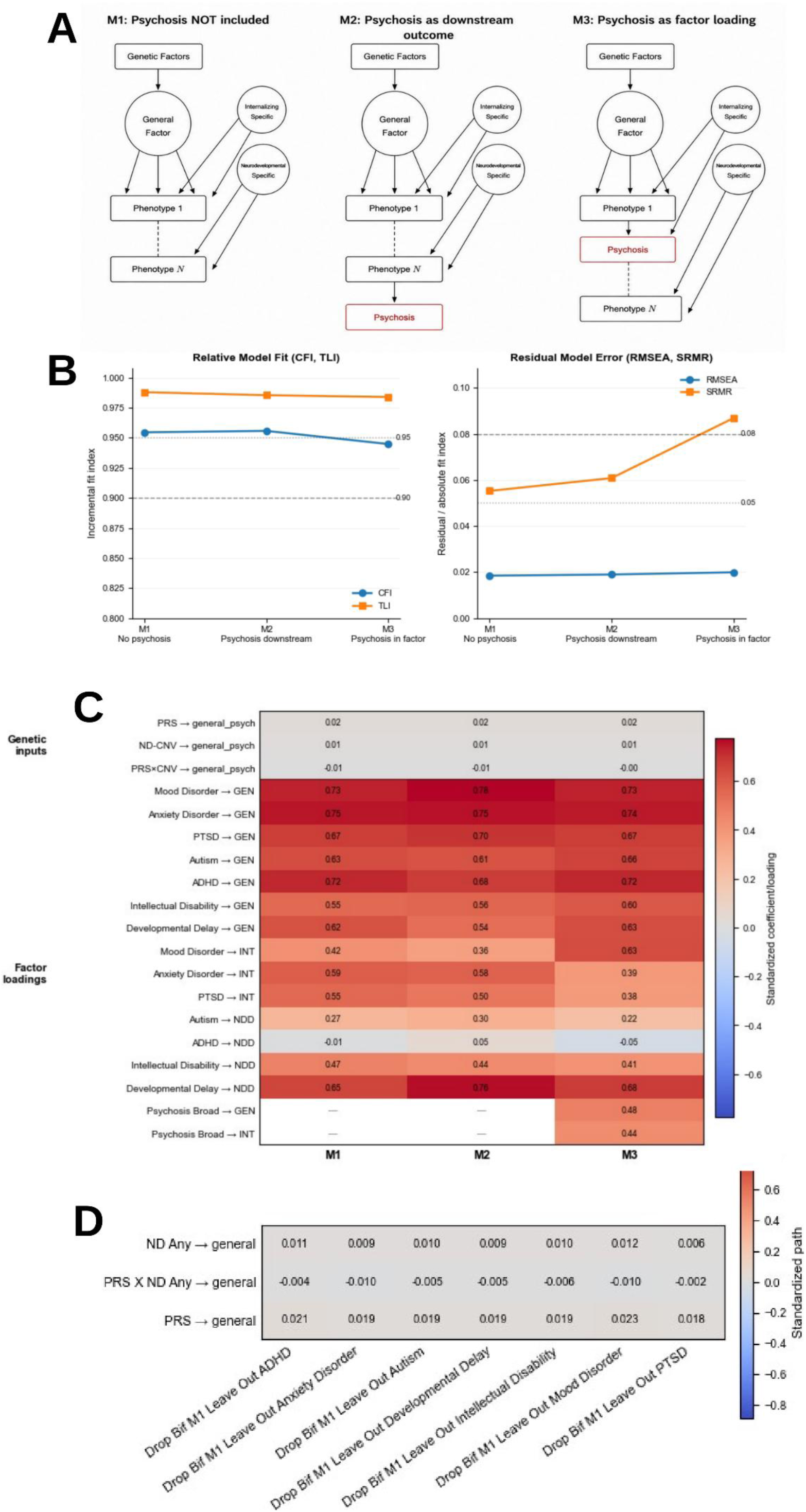
Covariance modeling of distributed neuropsychiatric phenotype organization. Structural equation modeling performed using the primary phenotype covariance matrix derived from the study cohort. **(A)** Schematic representation of bifactor structural equation modeling (SEM) psychosis embedding strategies. Model 1 (M1) excluded psychosis-spectrum phenotypes from the latent factor structure, Model 2 (M2) modeled psychosis as a downstream manifestation of the general covariance factor, and Model 3 (M3) incorporated psychosis directly as a factor-loading phenotype within the latent structure. All models incorporated schizophrenia polygenic risk scores (PRS), recurrent ND-CNV carrier status, and PRS × ND-CNV interaction effects. **(B)** Comparative model fit and residual error estimates across bifactor SEM psychosis embedding strategies. Panels summarize comparative fit index (CFI), Tucker-Lewis index (TLI), root mean square error of approximation (RMSEA), and standardized root mean square residual (SRMR) across M1-M3 frameworks. Lower RMSEA and SRMR indicate improved residual fit. **(C)** Heatmap summarizing standardized path coefficients and factor loadings across bifactor SEM psychosis embedding strategies. Columns correspond to M1-M3 frameworks, while rows summarize schizophrenia PRS, recurrent ND-CNV carrier status, PRS × ND-CNV interaction effects, latent-factor path estimates, and phenotype-level factor loadings. Color intensity reflects standardized effect magnitude and direction. **(D)** Leave-one-indicator-out (LOO) sensitivity analyses across bifactor SEM frameworks. Heatmaps summarize relative stability of covariance structure and standardized genetic path coefficients following sequential removal of individual phenotypic indicators from the bifactor structure.

Using the bifactor specification selected for downstream analyses, we next evaluated three alternative psychosis embedding strategies (**Figure 3A**). Models treating psychosis as external to (M1), or downstream of (M2), the latent factor structure revealed improved fit relative to models incorporating psychosis directly into the latent factor architecture (M3). M1 showed CFI = 0.955, TLI = 0.988, RMSEA = 0.018 (95% CI 0.018–0.019), and SRMR = 0.055, while M2 showed CFI = 0.956, TLI = 0.987, RMSEA = 0.019 (95% CI 0.019–0.020), and SRMR = 0.061. In contrast, M3 showed CFI = 0.945, TLI = 0.986, RMSEA = 0.020 (95% CI 0.020–0.021), and SRMR = 0.087 (**Figure 3B**).

Across SEM configurations, standardized path coefficients were robust and consistently positive for schizophrenia PRS and recurrent ND-CNV carrier status (β ≈ 0.019 and β ≈ 0.009, respectively), whereas PRS × ND-CNV interaction effects remained near zero (β ≈ −0.005). Similar parameter estimates were observed across all three model specifications (**Figure 3C**). Within the psychosis-downstream model (M2), the general factor was strongly associated with psychosis-spectrum phenotypes (β = 0.881). In contrast, direct paths from schizophrenia PRS (β = 0.006), recurrent ND-CNV carrier status (β = 0.005), and the PRS × ND-CNV interaction (β = 0.011) to psychosis were comparatively small after accounting for the general factor.

Additional sensitivity analyses similarly showed substantial stability of the primary findings (**Figure 3D**). Sequential leave-one-indicator-out analyses preserved the overall covariance structure and genetic path estimates, while reduced-factor models excluding the highly prevalent mood and anxiety phenotypes altered global fit characteristics without changing the principal structural relationships (**Figure S6C**). Within the reduced psychosis-downstream model, the general covariance factor continued to strongly predict psychosis-spectrum phenotypes (β = 0.892) (**Figure S6D**). Predictor-specific SEM sensitivity analyses produced similar results. Models including only recurrent ND-CNV carrier status or only schizophrenia PRS again identified the psychosis-downstream model (M2) as the best-fitting covariance structure (**Figure S7**).

## DISCUSSION

This study extends prior work on schizophrenia-associated genetic pleiotropy by jointly characterizing common polygenic liability and structural variation overlapping recurrent neurodevelopmental copy-number variant (ND-CNV) loci across a broad, longitudinal EHR-derived neuropsychiatric phenotype space. In the large, ancestrally diverse *All of Us* Research Program ^54^ cohort, schizophrenia polygenic risk score (PRS) and recurrent ND-CNV carrier status each showed distributed associations extending beyond schizophrenia diagnosis alone, while deletion- and duplication-overlapping carrier groups exhibited distinct aggregate association profiles. PRS × ND-CNV interaction estimates provided little evidence of widespread multiplicative interaction, suggesting largely independent statistical contributions at the resolution of these analyses. Complementary dimensional and latent-variable analyses further showed that these distributed associations could be summarized within organized patterns of covariance across psychiatric and neurodevelopmental phenotypes. Together, these findings demonstrate that associations with distinct forms of schizophrenia-associated genetic liability remain detectable across multiple representations of real-world longitudinal EHR phenotypes, providing a population-scale framework for examining how common and structural genomic variation are expressed across the broader neuropsychiatric phenotype space.

### Common and structural genetic risk in a shared phenotype space

Schizophrenia PRS and structural variation overlapping recurrent ND-CNV loci represent markedly different forms of genetic variation. Schizophrenia PRS captures the aggregate contribution of many common variants of individually small effect ^7^, whereas variants within ND-CNV loci capture structural alterations with substantially larger individual effects and stronger evolutionary constraint^19,45^. Despite these differences in genetic architecture, both forms of genetic liability showed associations extending across psychiatric, cognitive, and neurodevelopmental phenotypes in the present study. This convergence at the level of phenotype suggests that distinct forms of genomic liability can be expressed across partially overlapping dimensions of neuropsychiatric susceptibility^4,15,19,29,31,42^.

These findings are consistent with an expanding body of evidence that genetic liability associated with schizophrenia is not expressed exclusively through schizophrenia or psychosis^42,43,61,62^. Population-based studies of schizophrenia PRS have demonstrated associations across psychiatric, cognitive, and developmental traits ^63–65^, while structural variation at recurrent ND-CNV loci has independently been associated with pleiotropic risk across numerous neurodevelopmental and psychiatric conditions^23,28–30,32,44^. The present study brings these previously parallel observations into a common analytic framework by evaluating polygenic liability and structural variation across the same longitudinal EHR-derived phenotype space. Their distributed associations were therefore observable within the same population and phenotype definitions, allowing similarities and differences in their aggregate patterns of clinical expression to be evaluated directly rather than inferred across separate studies.

Importantly, phenotypic convergence does not imply that common polygenic liability and structural variation act through the same biological mechanisms. Similar or overlapping clinical outcomes could arise from shared, partially shared, or distinct molecular, cellular, developmental, and circuit-level processes^1,43,66–68^. The present analyses establish convergence at the level of distributed phenotype association but do not resolve where, or whether, the underlying biological pathways converge.

Identifying those intermediate mechanisms represents an important next step toward understanding how distinct forms of genomic liability ultimately contribute to overlapping neuropsychiatric outcomes.

### Shared and divergent architectures of deletion- and duplication-overlapping ND-CNVs

ND-CNV carrier groups exhibited distinct aggregate association profiles, with deletion-overlapping carriers generally showing larger and more consistently positive estimates than duplication-overlapping carriers, particularly across psychosis-spectrum and neurodevelopmental phenotypes. These differences suggest that pooling deletion- and duplication-overlapping events may obscure direction-specific heterogeneity in the expression of schizophrenia-associated structural genomic liability. However, separation by event type provides only a coarse representation of structural genomic variation. The substantial overlap that remained across deletion- and duplication-associated phenotype profiles indicates that these classes are neither phenotypically discrete nor uniformly divergent. Because the present analyses aggregated variants across recurrent loci rather than comparing reciprocal events within individual genomic regions, differences between these groups should not be interpreted as evidence for a uniform deletion-versus-duplication effect^55,57,69^.

This mixture of shared and direction-specific associations is consistent with the complex phenotypic consequences of gene dosage variation. Reciprocal copy-number changes affecting the same genomic interval can produce strongly asymmetric phenotypes^16,70–74^, but dosage gains and losses can also share neurodevelopmental manifestations. Their consequences depend not only on the direction of copy-number change but on the genes and regulatory elements affected ^75–79^, dosage sensitivity, genomic context, and the downstream coordination of interacting biological systems.

Classic examples include *PMP22* duplication and deletion, which cause Charcot-Marie-Tooth disease type 1A and hereditary neuropathy with liability to pressure palsies, respectively^80–82^ , and reciprocal 17p11.2 dosage changes involving *RAI1*, which produce Smith–Magenis and Potocki–Lupski syndromes^83,84^ . Reciprocal 16p11.2 CNVs further illustrate this complexity, producing mirror effects for some traits, including body mass and head circumference, while sharing other neurodevelopmental manifestations^85,86^. Thus, reciprocal dosage changes can produce divergent effects along some phenotypic dimensions while converging along others.

This distinction is particularly relevant to interpreting the distributed schizophrenia- and psychosis-spectrum associations observed here. The overlap between deletion- and duplication-associated profiles should not be viewed simply as residual heterogeneity that would disappear with more refined classification; rather, the coexistence of shared and direction-specific associations may itself reflect the pleiotropic expression of structural genomic liability across the neuropsychiatric phenotype space. Broad deletion and duplication categories collapse substantial heterogeneity in locus, genomic content, and dosage sensitivity, while distinct structural perturbations may nevertheless converge on overlapping psychosis-spectrum and neurodevelopmental outcomes. The replication of this mixed pattern in the ancestrally diverse *All of Us* cohort therefore reinforces a central feature of schizophrenia-associated structural genomic risk: its clinical expression is neither specific to schizophrenia nor reducible to a simple deletion-versus-duplication distinction. Progressively more informative models will require moving beyond both pooled carrier status and binary event classification toward locus- and dosage-sensitive representations capable of resolving the shared and divergent pathways through which structural genomic variation contributes to schizophrenia and related neurodevelopmental phenotypes^28,29,87,88^.

### Largely independent contributions of polygenic and structural genomic liability

Across phenotype-level, dimensional, and latent-variable analyses, schizophrenia PRS and ND-CNV carrier status showed largely independent statistical associations with neuropsychiatric phenotypes, with little evidence of widespread multiplicative interaction. Psychosis-spectrum prevalence increased across the PRS distribution among both carriers and non-carriers, while the carrier-associated elevation remained relatively stable across PRS deciles. Together, these findings suggest that structural variation overlapping recurrent ND-CNV loci does not uniformly substitute for common polygenic liability, but instead contributes additional neuropsychiatric risk across the range of schizophrenia polygenic liability represented in this cohort.

This pattern is consistent with an increasingly nuanced literature on the joint contributions of common and structural genomic variation to schizophrenia risk. Individuals with schizophrenia who carry established risk CNVs continue to show enrichment for common schizophrenia-associated alleles, indicating that structural genomic risk does not define a genetically isolated subgroup^48^. Conversely, some have found lower schizophrenia PRS among affected carriers of higher-impact CNVs than among affected non-carriers with schizophrenia, consistent with liability-threshold models in which greater structural genomic liability reduces the additional polygenic burden associated with disease^89^. Studies of 22q11.2 deletion syndrome further demonstrate that common polygenic background can modify schizophrenia penetrance and related cognitive and clinical phenotypes even in the presence of a high-impact structural variant^47,90^ . Together, these findings suggest that common and structural genomic liability can contribute jointly to schizophrenia while their quantitative relationship varies across loci and populations.

The near-null interaction estimates observed here therefore provide little evidence for widespread departure from multiplicative joint effects at the aggregate level, but should not be interpreted as demonstrating biological independence or excluding locus- or phenotype-specific interactions. The recurrent ND-CNV framework combines structurally and biologically heterogeneous loci, and statistical power for interaction testing was substantially lower than for main-effect analyses. The present findings are therefore most consistent with detectable contributions from both polygenic and structural genomic liability across the broader neuropsychiatric phenotype space, while leaving open how those contributions combine within individual recurrent CNVs ^42,47,63,91^.

### Covariance modeling complements phenotype-level association analyses

Dimensional and latent-variable analyses provided complementary representations of the distributed phenotype-level associations observed for schizophrenia PRS and recurrent ND-CNV carrier status. Because psychiatric and neurodevelopmental phenotypes are substantially correlated, modeling their covariance allowed us to examine whether the distributed genetic associations observed across individual outcomes could also be represented within broader dimensions of shared phenotypic variation. Among the covariance structures tested, bifactor models showed the most favorable overall pattern of fit and residual error and were therefore carried forward for structural equation modeling.

Across alternative SEM specifications, models representing psychosis separately from, or in relation to, the broader latent covariance structure were better supported than a model incorporating psychosis directly as an indicator of that structure. These findings describe the statistical organization of the EHR-derived phenotypes in the present cohort rather than establishing causal relationships, biological hierarchy, or a particular psychiatric nosology ^92,93^.

This distinction is particularly relevant to schizophrenia-associated genetic liability. The distributed associations observed for schizophrenia PRS and ND-CNV carrier status were not confined to psychosis-spectrum diagnoses but extended across phenotypes contributing to broader dimensions of psychiatric and neurodevelopmental covariance. At the same time, psychosis was not most favorably represented as simply another indicator within that shared latent structure. Together, these observations suggest that studying schizophrenia-associated genetic liability across a broader phenotype space can capture dimensions of its clinical expression that are not apparent from schizophrenia diagnosis alone, while preserving psychosis as a distinct component of that organization^43,62,67,94,95^. Importantly, the latent structures identified here provide statistical summaries of phenotypic covariance rather than mechanistic explanations for why genetic liability is distributed across these outcomes.

The next challenge is therefore to determine which intermediate biological and cognitive processes connect genetic liability and shared dimensions of neuropsychiatric variation to the clinical phenomena we recognize as psychosis. Many of these intermediate processes remain unmeasured when psychiatric phenotypes are defined primarily through observable behavior, patient-reported experience, and clinical diagnosis^96,97^. Molecular networks, cell-type-specific processes, neurodevelopmental programs, and neural circuits represent candidate levels of organization through which distinct forms of genomic variation could ultimately contribute to overlapping clinical phenotypes^97,98^. Connecting these mechanistically proximal processes to the statistical phenotype structures observed here will require data that directly measure them; neither individual diagnostic associations nor latent covariance models alone can establish those biological relationships.

### Limitations and Future Directions

Several limitations should be considered. First, EHR-derived psychiatric phenotypes reflect clinical recognition, healthcare utilization, and coding practices in addition to underlying disease; adjustment for utilization and observation duration cannot eliminate differential ascertainment. Similarly, the RDoC-informed domains represent groupings of EHR diagnoses rather than direct measurement of RDoC constructs. Schizophrenia PRS performance also remains imperfect across diverse populations despite ancestry-aware harmonization, and the reduced 2,200-variant PRS may capture less polygenic information than the complete source score.

Second, recurrent ND-CNV carrier status was defined by structural-variant overlap with curated genomic intervals and does not constitute clinical adjudication of pathogenicity or resolve inheritance or canonical breakpoints. Pooling across recurrent loci combines substantial heterogeneity in genomic content and dosage sensitivity, while deletion- and duplication-overlapping groups remain broad, non-mutually exclusive categories rather than direct reciprocal comparisons. The framework was also not designed to estimate cumulative CNV burden. Future larger studies will be needed to resolve locus- and dosage-specific associations.

Third, the smaller number of ND-CNV carriers limited power for interaction analyses. Near-null PRS × ND-CNV interaction estimates therefore do not establish biological independence or exclude phenotype- or locus-specific interactions. Likewise, CFA and SEM summarize covariance conditional on the phenotypes and models examined and do not establish causal or biological relationships.

Despite these limitations, schizophrenia polygenic liability and structural variation overlapping recurrent ND-CNV loci demonstrated broadly distributed neuropsychiatric associations, with little evidence of widespread multiplicative interaction and both shared and direction-specific deletion- and duplication-associated profiles. Integrating increasingly resolved genomic variation with longitudinal phenotyping and direct measures of molecular, cellular, cognitive, and neural processes will be important for understanding how genomic liability ultimately contributes to psychosis and related neuropsychiatric phenotypes.

## METHODS

### Study cohort and data resource

All analyses were performed using the National Institutes of Health (NIH) *All of Us* Research Program^54^ Controlled Tier version 8 (CDRv8) dataset within the *All of Us* Researcher Workbench cloud environment^99^. The *All of Us* Research Program is a longitudinal United States precision medicine initiative integrating electronic health records (EHR), whole-genome sequencing (WGS)^100^, genotyping array data, survey instruments, physical measurements, and biospecimen-linked molecular resources across a large ancestrally diverse participant cohort. Controlled Tier analyses were conducted under approved researcher access protocols within the secure cloud-native Researcher Workbench infrastructure. The primary analytic cohort was generated through harmonization of recurrent neurodevelopmental copy-number variant (ND-CNV) carrier status, schizophrenia polygenic risk scores (PRS), EHR-derived psychiatric phenotypes, demographic covariates, healthcare-utilization variables, and ancestry principal components. Participants lacking harmonized genomic or longitudinal phenotype data required for downstream modeling were excluded from integrated analyses. The final analytic cohort included 71,992 participants, including 2,115 recurrent ND-CNV carriers.

### Curated recurrent ND-CNV framework

Throughout this study, the term “recurrent ND-CNV” refers to structural variants overlapping curated recurrent genomic intervals that have been repeatedly implicated in neurodevelopmental and psychiatric disorders. The term does not distinguish between inherited and de novo events, both of which may occur at these loci. Recurrent neurodevelopmental copy-number variant intervals were curated from established psychiatric and neurodevelopmental genetics literature, including recurrent loci previously implicated in schizophrenia, autism spectrum disorder, intellectual disability, developmental delay, epilepsy, and related neuropsychiatric phenotypes. Core interval resources were assembled from prior large-scale CNV studies^23,28,29,44^, followed by harmonization against ClinGen Dosage Sensitivity Map^101^ resources, Online Mendelian Inheritance in Man (OMIM)^102^, and ClinVar^103^. All intervals were harmonized to GRCh38 genomic coordinates using UCSC liftOver^104^ where required. Primary analyses used a reciprocal-overlap threshold of ≥30% (ro30) for participant-level carrier assignment. Additional overlap thresholds were generated during exploratory sensitivity analyses but were not used for primary downstream modeling. Curated intervals were additionally intersected against multiple schizophrenia-relevant and neurodevelopmental annotation resources, including loss-of-function intolerant genes from the Genome Aggregation Database (gnomAD) LOEUF framework^58^, Simons Foundation Autism Research Initiative (SFARI) autism-associated genes^59^, Psychiatric Genomics Consortium (PGC) schizophrenia-prioritized genes^7^, schizophrenia coding-gene resources, and schizophrenia-associated eQTL-linked genes.

### Structural variant processing and participant-level carrier assignment

Structural variant processing was performed using whole-genome sequencing structural variant resources available through the *All of Us* short-read sequencing pipeline^100^. Genomic analyses were conducted using Hail (version 0.2), Apache Spark, Python (version 3.10), and BigQuery-integrated workflows within the Researcher Workbench environment. Structural variants were intersected against curated recurrent ND-CNV intervals using reciprocal overlap criteria. Inspection of raw structural variant calls revealed substantial fragmentation heterogeneity across chromosomes and recurrent loci, resulting in multiple overlapping calls for individual participant-level events. Participant-level carrier definitions were therefore selected to avoid inflation of carrier status estimates arising from locus-specific differences in structural variant call fragmentation. This approach was selected because the biological unit of interest was recurrent locus carrier status rather than the number of fragmented structural variant calls generated during variant detection.

Participants carrying one or more structural variants satisfying overlap criteria within a curated interval were assigned binary indicators for pooled ND-CNV carrier status, deletion-overlapping ND-CNV carrier status (DEL), and duplication-overlapping ND-CNV carrier status (DUP). Because participant-level carrier assignments were generated independently for deletion-overlapping and duplication-overlapping events, individuals carrying both classes of recurrent ND-CNVs could contribute to both carrier groups. Chromosome-level row-density analyses were additionally performed to evaluate fragmentation-related heterogeneity across deletion and duplication calls and to justify participant-level rather than row-level structural variant modeling.

### Schizophrenia polygenic risk score generation

Schizophrenia PRS were generated using the AoUPRS software package (v0.2.6; Khattab, Scripps Research) within the *All of Us* Researcher Workbench, using PGS Catalog score PGS002785 and *All of Us* v8 WGS VDS resources. AoUPRS integrates PGS Catalog harmonization, ancestry-aware normalization, scalable cloud-native processing, and downstream phenotype harmonization directly within the *All of Us* computational environment. To facilitate computationally scalable implementation within the *All of Us* cloud environment, a reduced PRS was generated by selecting the highest-weighted variants from each autosome, resulting in a harmonized set of 2,200 variants (100 variants per autosome). The reduced score was subsequently harmonized, ancestry-adjusted, residualized, and standardized using the AoUPRS workflow to generate the exposure variable *prs_scz_sem_z*.

Validation analyses evaluated preservation of genome-wide effect-weight architecture, positive and negative weight balance, genomic distribution, and the expected relationship between schizophrenia polygenic burden and psychosis-spectrum phenotypes (**Figures S2–S3**). Population structure correction incorporated ancestry principal components (PC1–PC5) in all downstream regression and latent-variable analyses.

### Psychiatric phenotype derivation and RDoC mapping

Psychiatric and neurodevelopmental phenotypes were derived from OMOP^105^ Common Data Model concept mappings available through the *All of Us* EHR infrastructure. Phenotype definitions incorporated curated OMOP concept sets spanning psychosis-spectrum disorders, schizophrenia-spectrum disorders, schizoaffective phenotypes, mood disorders, anxiety disorders, post-traumatic stress disorder (PTSD), autism spectrum disorder, attention-deficit/hyperactivity disorder (ADHD), intellectual disability, developmental delay, epilepsy, and related psychiatric and neurodevelopmental conditions. Phenotypes were additionally mapped to Research Domain Criteria^97,106^ (RDoC)-informed dimensional frameworks using a curated psychiatric phenotype dictionary integrating categorical psychiatric diagnoses with broader neurobehavioral domains. Primary RDoC domains included cognitive systems, negative valence systems, positive valence systems, social processes, arousal/regulatory systems, and sensorimotor systems. Binary participant-level phenotype indicators were generated following longitudinal collapse of EHR-linked diagnostic coding across available observation periods. Individual diagnostic phenotypes were permitted to contribute to multiple RDoC domains when conceptually appropriate, reflecting the multidimensional nature of psychiatric symptomatology.

### Regression modeling framework

Recurrent ND-CNV carrier status was modeled as a binary participant-level indicator reflecting the presence of one or more curated recurrent ND-CNV intervals meeting reciprocal-overlap criteria. Primary association analyses evaluated pooled ND-CNV carrier status, deletion-overlapping ND-CNV carrier status, and duplication-overlapping ND-CNV carrier status across psychosis-spectrum and broader psychiatric phenotypes using logistic regression models. Secondary analyses subsequently incorporated schizophrenia PRS and PRS × ND-CNV interaction terms. Interaction models evaluated departure from multiplicative effects between schizophrenia PRS and recurrent ND-CNV carrier status. Because schizophrenia PRS was standardized before modeling, PRS main-effect odds ratios represent the change in outcome odds per 1-SD increase in PRS. ND-CNV main-effect odds ratios compare carriers with non-carriers at the mean PRS value, and exponentiated PRS × ND-CNV interaction coefficients represent the ratio of the per-SD PRS odds ratios between carriers and non-carriers.

Primary regression models were adjusted for age, sex, ancestry principal components (PC1-PC5), healthcare-utilization burden, and longitudinal observation duration. Healthcare-utilization adjustment incorporated log-transformed number of unique clinical encounters, while longitudinal observation adjustment incorporated total EHR-linked observation time. Phenotype-level analyses estimated associations for individual diagnoses, whereas dimensional analyses evaluated whether associations converged across broader RDoC-informed neurobehavioral domains. Odds ratios, confidence intervals, and p-values were extracted from downstream regression outputs. Given the substantial clinical, definitional, and statistical dependence among the modeled neuropsychiatric phenotypes, primary results are reported using effect estimates, 95% confidence intervals, and nominal two-sided *p* values rather than interpreting individual associations according to a single multiple-testing threshold. Benjamini–Hochberg false discovery rate (FDR)–adjusted *q* values were additionally calculated across phenotype-level analyses and are provided in the Supplementary Materials as a complementary measure of multiplicity.

Sensitivity analyses evaluated robustness of phenotype-level association architecture across alternate covariate structures, demographic perturbations, healthcare-utilization perturbations, ancestry principal component perturbations, and observation-time sensitivity models. Deletion-overlapping and duplication-overlapping carrier sensitivity analyses additionally compared psychiatric architectures associated with deletion-overlapping and duplication-overlapping ND-CNV carrier status across both phenotype-level and dimensional analyses. Results from phenotype-level and dimensional regression analyses informed subsequent latent-variable modeling designed to evaluate shared covariance structure across psychiatric phenotypes.

### Confirmatory factor analysis and structural equation modeling

Confirmatory factor analysis (CFA) and structural equation modeling (SEM) were performed in R using the lavaan^92^ package version 0.6-21. Because modeled psychiatric and neurodevelopmental indicators were EHR-derived binary variables, models were estimated using weighted least squares with mean and variance adjustment (WLSMV), reported by lavaan as diagonally weighted least squares (DWLS), under a liability-threshold framework with theta parameterization. Phenotypic indicators were modeled as ordered binary variables, and missing data were handled using pairwise present estimation implemented within lavaan.

Model evaluation incorporated comparative fit index (CFI), Tucker-Lewis index (TLI), root mean square error of approximation (RMSEA), standardized root mean square residual (SRMR), chi-square statistics, and model degrees of freedom. Initial CFA analyses compared one-factor, correlated two-factor, and bifactor latent structures. Bifactor models demonstrated the most favorable balance of relative fit and residual error metrics and were therefore selected for downstream SEM analyses.

Structural equation modeling integrated schizophrenia PRS, ND-CNV carrier status, and PRS × ND-CNV interaction effects across three alternative psychosis embedding strategies (**Figure 3A**). Model 1 (M1) excluded psychosis-spectrum phenotypes from the latent factor structure, Model 2 (M2) modeled psychosis downstream of the broader latent psychiatric/developmental liability factor, and Model 3 (M3) incorporated psychosis directly as a factor-loading phenotype within the latent structure.

To evaluate whether latent-model architecture depended upon simultaneous modeling of common and rare genetic risk, the three SEM formulations were additionally re-estimated using schizophrenia PRS alone and recurrent ND-CNV carrier status alone as the sole genetic predictor. Comparative model-fit metrics across these reduced predictor specifications were evaluated to determine whether preferred psychosis embedding strategies were robust to genetic predictor selection.

Additional SEM sensitivity analyses evaluated the robustness of latent architecture across alternative phenotype and predictor specifications. Leave-one-indicator-out (LOO) analyses sequentially removed individual phenotypic indicators from the bifactor structure to evaluate stability of latent-factor organization. Reduced-factor analyses excluded mood and anxiety phenotypes to determine whether highly prevalent affective disorders disproportionately influenced latent covariance structure. Finally, PRS-only and ND-CNV-only SEMs evaluated whether preferred psychosis embedding strategies remained stable when common and rare genetic liability were modeled independently.

### Statistical environment and software

Primary genomic processing and recurrent ND-CNV identification were performed within the *All of Us* Researcher Workbench (Controlled Tier v8) using Hail, Apache Spark, Python, and BigQuery-integrated workflows. Phenotype extraction, cohort assembly, and regression analyses were conducted using Python (version 3.10) with pandas, NumPy, SciPy, and statsmodels. Statistical analyses requiring latent-variable modeling were performed locally in R (version 4.3) using the lavaan package (version 0.6-21).

Data visualization incorporated matplotlib, seaborn, and ggplot2 together with custom Python and R plotting scripts developed for phenotype-level association analyses, dimensional RDoC summaries, genomic interval visualization, structural equation modeling, and sensitivity analyses. All analyses were conducted within the secure *All of Us* Researcher Workbench environment unless otherwise noted.

## Data Availability

Additional supporting materials, including complete machine-readable result tables, figure source data, analysis notebooks, code, quality-control outputs, and extended model results, are available through the accompanying Open Science Framework repository.

https://osf.io/4c58y/overview?view_only=4b449a32ac17473d89471a32f3c6c904.

## Funding/Acknowledgements

We thank the participants in the NIH *All of Us* Research Program for their trust in science and willingness to contribute to accelerating translational medicine. We also thank the AoU Research Program support for generating and providing the infrastructure and maintaining the dataset, as well as the integrity of the data for all participants, that made this work possible. The training for AMW and AD was funded by T32GM113896 / T32GM145432 (AMW & AD); F30MH134482 (AMW); F30 AG087698 / T32AG082661 (AD). AR is supported by Faculty STARs Program, P30AG066546 (AR).

## Notes

### Competing Interest Statement

The authors have declared no competing interest.

### Author Declarations

NIH All of Us Research Program

