## Supplement for "Schizophrenia-associated polygenic liability and structural genomic risk demonstrate broadly distributed neuropsychiatric associations in the *All of Us* Research Program"

**Table of Contents**


| Section | Title | Page |
|---|---|---|
| I | Supplemental Methods | 2 |
| II | Code Description for AoU ND-CNV, Schizophrenia PRS, and Logistic Regression Pipeline | 12 |
| III | External notebooks and locally executed analysis packages | 24 |
| IV | Code Description for the Lavaan CFA/SEM Pipeline | 31 |
| V | Supplemental Figures | 43 |
| VI | External Supplemental Tables | 55 |

### I. Supplemental Methods

#### Computational environment and genomic infrastructure

All participant-level genomic, phenotype, and regression analyses were conducted within the National Institutes of Health *All of Us* Research Program Researcher Workbench Controlled Tier version 8 (CDRv8) environment. Analyses relying exclusively on aggregate, de-identified, or publicly available resources, including selected genomic annotation and latent-variable analyses, were conducted locally as described below. The *All of Us* Research Program provides cloud-native access to harmonized genomic, electronic health record, survey, and biospecimen-linked participant data within a secure analysis infrastructure. Structural variant processing and genomic analyses were performed using Hail version 0.2, Apache Spark distributed computing infrastructure, Google BigQuery SQL, and Python version 3.10 within the Researcher Workbench environment.

Whole-genome sequencing resources were derived from the *All of Us* short-read sequencing pipeline. Structural variant resources incorporated population-scale short-read WGS-derived structural variant calls available through Controlled Tier access.

#### Recurrent ND-CNV interval curation and harmonization

Curated recurrent genomic intervals containing neurodevelopmental copy-number variant (ND-CNV)-associated loci were assembled from established neurodevelopmental and psychiatric genetics literature, as previously described by our group (Wells et al., 2026. *Medrxiv*: https://doi.org/10.64898/2026.09.13.26362959). Throughout this supplement, “recurrent ND-CNV” refers to structural variants overlapping curated recurrent genomic intervals that have been repeatedly implicated in neurodevelopmental and psychiatric disorders. This terminology does not establish that participant-carried events arose independently or de novo and does not distinguish inherited from de novo variation.

Primary interval resources incorporated recurrent loci previously implicated in schizophrenia, autism spectrum disorder, intellectual disability, developmental delay, epilepsy, and related neuropsychiatric phenotypes from prior large-scale CNV studies. Following exclusion of the chromosome X interval, the curated framework contained 96 autosomal direction-specific deletion and duplication locus definitions corresponding to 51 unique autosomal genomic intervals.

Intervals were cross-referenced against ClinGen Dosage Sensitivity Map resources, Online Mendelian Inheritance in Man (OMIM), ClinVar, and additional recurrent neurodevelopmental CNV references. All intervals were harmonized to GRCh38 genomic coordinates using UCSC liftOver where required. Primary downstream analyses used reciprocal overlap thresholds ≥30% (ro30) for participant-level carrier assignment.

#### Structural variant intersection and participant-level carrier assignment

Structural variant interval intersection analyses were performed using Hail-based genomic workflows integrated with Spark-distributed processing. Structural variants were intersected against curated recurrent ND-CNV intervals using reciprocal overlap criteria.

Because raw structural variant resources demonstrated substantial row-level fragmentation heterogeneity across chromosomes and loci, participant-level interval-collapsed carrier definitions were used throughout downstream analyses rather than row-level structural variant counts. This approach minimized inflation of structural variant carrier status resulting from segmentation-related fragmentation of large structural events.

Participants carrying one or more structural variants satisfying overlap criteria within a curated interval were assigned binary indicators for: (i) pooled ND-CNV carrier status, (ii) deletion-specific carrier status (DEL), (iii) duplication-specific carrier status (DUP). Deletion-overlapping and duplication-overlapping classifications were generated independently and were not mutually exclusive; participants carrying both event classes contributed to both direction-specific groups. Chromosome-level fragmentation analyses were additionally performed to evaluate row-density heterogeneity across deletion and duplication calls.

**Broad schizophrenia GWAS discovery gene overlap analysis**

To further benchmark the curated recurrent ND-CNV framework against the broader schizophrenia GWAS landscape, recurrent ND-CNV intervals were additionally intersected with the broad FINEMAP-derived schizophrenia discovery gene set reported by Trubetskoy *et al.* This secondary analysis was performed outside the primary All of Us Researcher Workbench pipeline using locally executed Python workflows because it relied exclusively on publicly available genomic annotation resources and did not require access to participant-level data. Protein-coding genes from the published FINEMAP discovery set were mapped to GENCODE v38 (GRCh38) gene coordinates prior to interval intersection. Gene-level overlap was quantified by intersecting curated recurrent ND-CNV intervals with annotated gene coordinates, followed by aggregation of locus-level overlap counts, interval-normalized gene density (genes/Mb), and participant-independent overlap summaries. Resulting overlap statistics were used to generate the locus-level benchmarking visualization presented in **Figure 1B**.

**Genomic annotation resource integration**

Curated recurrent ND-CNV intervals were intersected against multiple schizophrenia-relevant and neurodevelopmental genomic annotation resources. Loss-of-function intolerant genes were derived from the Genome Aggregation Database (gnomAD) loss-of-function observed/expected upper bound fraction (LOEUF) framework. Autism-associated genes were derived from the Simons Foundation Autism Research Initiative (SFARI) Gene database. Schizophrenia-associated annotation resources incorporated Psychiatric Genomics Consortium (PGC)-prioritized schizophrenia genes, schizophrenia coding genes, and schizophrenia-associated expression quantitative trait loci (eQTL)-linked genes derived from published schizophrenia GWAS integration studies. Gene-level overlap analyses quantified annotation overlap across recurrent ND-CNV intervals (**Figure 1C**) and participant-level carrier groups (**Figure 1D**).

#### Polygenic risk score generation using AoUPRS

Schizophrenia polygenic risk scores (PRS) were generated using AoUPRS, a scalable ancestry-aware PRS framework recently developed specifically for implementation within the *All of Us* Research Program cloud environment. AoUPRS integrates cloud-native genomic processing, PGS Catalog harmonization, ancestry-aware normalization, and downstream phenotype integration workflows optimized for the computational architecture of the Researcher Workbench ecosystem.

AoUPRS was selected because it currently represents the most robust and computationally feasible framework for large-scale PRS implementation within *All of Us*, where traditional local PRS workflows remain difficult to operationalize efficiently across population-scale genomic datasets.

Schizophrenia PRS was generated using AoUPRS version 0.2.6 and PGS Catalog score PGS002785. A computationally reduced score was constructed by retaining the 100 variants with the largest absolute weights from each autosome, yielding 2,200 variants. Downstream PRS harmonization incorporated ancestry-aware residualization and normalization procedures. Population structure correction incorporated ancestry principal components PC1-PC5 throughout downstream regression and latent-variable analyses. The primary harmonized schizophrenia PRS exposure variable used across integrated analyses was prs_scz_sem_z. Residualized schizophrenia PRS distributions were evaluated across ND-CNV carrier and non-carrier groups prior to downstream association analyses.

#### Psychiatric phenotype derivation and dimensional mapping

Psychiatric and neurodevelopmental phenotypes were derived from OMOP Common Data Model concept mappings available through the *All of Us* electronic health record infrastructure. Curated OMOP concept sets incorporated psychosis-spectrum disorders, schizophrenia-spectrum disorders, schizoaffective phenotypes, hallucination and delusion phenotypes, mood disorders, anxiety disorders, post-traumatic stress disorder (PTSD), autism spectrum disorder, attention-deficit/hyperactivity disorder (ADHD), intellectual disability, developmental delay, epilepsy, and additional psychiatric and neurodevelopmental conditions.

Phenotypes were additionally integrated into a curated Research Domain Criteria (RDoC)-informed dimensional framework. RDoC mapping incorporated cognitive systems, negative valence systems, positive valence systems, social processes, arousal/regulatory systems, and sensorimotor systems. Binary participant-level phenotype indicators were generated following longitudinal collapse of diagnostic coding across all available EHR-linked observation periods.

### Covariate Definitions and Harmonization

Primary regression and latent-variable models incorporated demographic, ancestry, healthcare-utilization, and longitudinal ascertainment covariates derived from harmonized *All of Us* Research Program participant-level data resources.

Age was defined using the participant age at most recent EHR-linked observation (age_last_obs) available within the harmonized longitudinal dataset. Sex was derived from harmonized *All of Us* demographic variables using binary sex assignments available within the Controlled Tier participant tables. Population structure adjustment incorporated ancestry principal components PC1-PC5 derived from *All of Us* genomic ancestry inference pipelines. These principal components were included in all primary regression and SEM analyses to minimize residual confounding arising from continental ancestry structure and subtle population stratification.

Healthcare-utilization burden was modeled using the variable log1p_n_unique_visits, defined as the natural log-transformed count of unique EHR-linked clinical visits plus one. This variable was included to reduce ascertainment bias arising from differential healthcare-system interaction intensity across participants. Longitudinal ascertainment duration was modeled using observation_time_days, defined as the total number of days between earliest and most recent EHR-linked observations available for each participant. Observation duration was included to account for differential opportunity for psychiatric diagnosis capture across participants with varying lengths of longitudinal follow-up.

For schizophrenia polygenic risk score harmonization, ancestry-aware normalization and residualization procedures incorporated demographic covariates and ancestry principal components. The primary downstream harmonized schizophrenia PRS exposure variable (prs_scz_sem_z) represented the standardized residualized schizophrenia PRS used across regression and latent-variable analyses.

### Logistic regression analyses

Primary association analyses were performed using logistic regression models evaluating the relationship between recurrent neurodevelopmental copy-number variant (ND-CNV) carrier status and psychiatric phenotypes derived from longitudinal EHR-linked OMOP concept mappings. Initial primary models focused on pooled participant-level ND-CNV carrier status across psychosis-spectrum and broader psychiatric phenotypes. Direction-specific analyses subsequently evaluated deletion-specific (DEL) and duplication-specific (DUP) carrier status independently to assess potential differences between reciprocal structural variant classes.

Secondary analyses subsequently incorporated schizophrenia polygenic risk scores (PRS) and PRS × ND-CNV interaction terms to evaluate whether schizophrenia-associated common polygenic liability modified observed ND-CNV-associated phenotypic covariance structure. These analyses were motivated by the substantial overlap between recurrent ND-CNV loci and schizophrenia-relevant genomic annotation resources, including schizophrenia-prioritized genes, coding loci, and eQTL-linked schizophrenia-associated genes.

All primary logistic regression models incorporated demographic, ancestry, healthcare-utilization, and longitudinal ascertainment covariates. Specifically, models adjusted for age at most recent EHR-linked observation (age_last_obs), sex, ancestry principal components PC1-PC5, healthcare-utilization burden (log1p_n_unique_visits), and total longitudinal EHR-linked observation duration (observation_time_days). Inclusion of healthcare-utilization and observation-duration covariates was intended to reduce ascertainment bias arising from differential healthcare-system interaction intensity and unequal longitudinal opportunity for psychiatric diagnosis capture.

Primary downstream schizophrenia PRS analyses used the standardized ancestry-aware harmonized exposure variable prs_scz_sem_z generated through the AoUPRS framework. Interaction analyses incorporated multiplicative PRS × ND-CNV interaction terms to evaluate whether schizophrenia-associated common polygenic liability modified recurrent ND-CNV-associated psychiatric covariance structure beyond additive effects alone. Because the schizophrenia PRS exposure was standardized before modeling, PRS main-effect odds ratios represent the change in outcome odds per 1-SD increase in PRS. ND-CNV main-effect odds ratios compare carriers with non-carriers at the mean PRS value. Exponentiated PRS × ND-CNV interaction coefficients represent the ratio of the per-SD PRS odds ratios between carriers and non-carriers.

Phenotype-level analyses evaluated individual psychiatric and neurodevelopmental outcomes spanning psychosis-spectrum disorders, schizophrenia-spectrum disorders, mood disorders, anxiety disorders, PTSD, autism spectrum disorder, ADHD, developmental delay, intellectual disability, epilepsy, and related neuropsychiatric conditions. Additional dimensional analyses evaluated grouped Research Domain Criteria (RDoC)-informed psychiatric domains including cognitive systems, negative valence systems, positive valence systems, social processes, arousal/regulatory systems, and sensorimotor systems.

Odds ratios, 95% confidence intervals, standardized effect estimates, and p-values were extracted from downstream regression outputs. Phenotype-level association structures were subsequently visualized using effect-size-scaled dot plots, domain-level radar plots, and heatmap-based perturbation analyses to evaluate broader distributed psychiatric covariance structure across pooled ND-CNV carrier status, deletion-specific carrier status, duplication-specific carrier status, schizophrenia PRS, and PRS × ND-CNV interaction models.

#### Regression sensitivity analyses and covariate perturbation modeling

Sensitivity analyses evaluated robustness of phenotype-level association architecture across alternate covariate structures and model perturbations. These analyses incorporated demographic perturbations, ancestry principal component perturbations, healthcare-utilization perturbations, and observation-time sensitivity models. Direction-specific analyses additionally evaluated deletion-specific and duplication-specific psychiatric covariance structures across phenotype-level and domain-level models. Heatmap-based perturbation analyses summarized relative stability of phenotype-level association structure across alternate regression specifications.

**Confirmatory factor analysis (CFA)**

Because phenotype-level and RDoC-informed regression analyses demonstrated broad but structured pleiotropic association covariance structure spanning psychosis-spectrum, neurodevelopmental, cognitive, and internalizing phenotypes, confirmatory factor analysis (CFA) was subsequently performed to evaluate whether observed phenotype covariance patterns were consistent with broader latent psychiatric/developmental liability structures.

Confirmatory factor analysis (CFA) was performed in R using the lavaan package version 0.6-21. Because modeled psychiatric and neurodevelopmental indicators were EHR-derived binary variables, models were estimated using weighted least squares with mean and variance adjustment (WLSMV), reported by lavaan as diagonally weighted least squares (DWLS), under a liability-threshold framework with theta parameterization. Phenotypic indicators were modeled as ordered binary variables, and missing data were handled using pairwise present estimation implemented within lavaan.

Three primary latent structures were evaluated:

1. A one-factor model representing a single shared latent psychiatric/developmental liability factor underlying all modeled phenotypes.
2. A correlated two-factor model representing partially separable but correlated latent psychiatric domains.
3. A bifactor model incorporating both a dominant shared latent psychiatric/developmental liability factor and partially orthogonal domain-specific variance structure.

Observed phenotypic indicators incorporated psychosis-spectrum disorders, schizophrenia-spectrum phenotypes, mood disorders, anxiety disorders, PTSD, autism spectrum disorder, ADHD, developmental delay, intellectual disability, and related psychiatric and neurodevelopmental phenotypes derived from OMOP-mapped EHR concept definitions.

The one-factor framework was selected to evaluate whether psychiatric covariance structure could be parsimoniously explained through a single dominant latent psychiatric/developmental liability dimension. The correlated two-factor framework was selected to evaluate whether psychiatric covariance was better represented through partially separable but correlated latent dimensions. Finally, bifactor frameworks were selected to evaluate whether psychiatric covariance structure simultaneously reflected both a dominant shared liability dimension and partially distinct orthogonal variance components.

Model evaluation incorporated comparative fit index (CFI), Tucker-Lewis index (TLI), root mean square error of approximation (RMSEA), standardized root mean square residual (SRMR), chi-square statistics, and model degrees of freedom. Although all evaluated CFA structures demonstrated generally strong incremental fit characteristics, bifactor models demonstrated the lowest residual misfit estimates across RMSEA and SRMR relative to one-factor and correlated two-factor alternatives. Because the primary goal of downstream latent-variable modeling was to evaluate distributed psychiatric covariance structure while simultaneously preserving partially separable phenotype-level covariance structure, bifactor frameworks were selected for downstream structural equation modeling analyses.

**Structural equation modeling**

Structural equation modeling (SEM) analyses were performed using bifactor latent structures to integrate schizophrenia polygenic risk scores (PRS), recurrent neurodevelopmental copy-number variant (ND-CNV) carrier status, and PRS × ND-CNV interaction effects within broader latent psychiatric/developmental covariance structure.

SEM analyses were motivated by the observation that psychosis-spectrum phenotypes demonstrated strong and directionally coherent associations with both schizophrenia PRS and recurrent ND-CNV carrier status during primary phenotype-level regression analyses. Psychosis was evaluated under three alternative statistical placements: excluded from the latent factor structure, modeled downstream of the broader factor, or included directly as a factor-loading indicator.

All SEM analyses used bifactor latent structures selected from preceding CFA analyses. The bifactor framework incorporated both a dominant shared latent psychiatric/developmental liability factor and partially orthogonal phenotype-specific variance structure.

Observed phenotypic indicators incorporated psychosis-spectrum disorders, schizophrenia-spectrum phenotypes, mood disorders, anxiety disorders, PTSD, autism spectrum disorder, ADHD, developmental delay, intellectual disability, and related psychiatric and neurodevelopmental phenotypes derived from longitudinal OMOP-linked EHR concept mappings.

Three alternative SEM psychosis embedding strategies were evaluated:

1. Model 1 (M1) excluded psychosis-spectrum phenotypes from the latent factor structure entirely. This model was designed to evaluate whether schizophrenia-associated genetic predictors were associated with the broader latent psychiatric/developmental covariance structure independent of psychosis-spectrum phenotypes.
2. Model 2 (M2) modeled psychosis downstream of the broader latent psychiatric/developmental liability factor. This framework evaluated whether psychosis was statistically represented as an outcome predicted by the broader factor.

3. Model 3 (M3) incorporated psychosis directly as a factor-loading phenotype within the bifactor latent structure itself. This model evaluated whether psychosis behaved as an integrated central component of broader latent psychiatric/developmental covariance structure.

All SEM analyses incorporated standardized schizophrenia PRS exposure variables generated using the AoUPRS framework, participant-level recurrent ND-CNV carrier status variables, and multiplicative PRS × ND-CNV interaction terms.

Structural models estimated standardized path coefficients linking schizophrenia PRS, recurrent ND-CNV carrier status, and interaction effects to latent psychiatric/developmental liability structure and downstream psychosis-spectrum phenotypes. SEM was performed in R using the lavaan package version 0.6-21. Because modeled psychiatric and neurodevelopmental indicators were EHR-derived binary variables, models were estimated using weighted least squares with mean and variance adjustment (WLSMV), reported by lavaan as diagonally weighted least squares (DWLS), under a liability-threshold framework with theta parameterization. Phenotypic indicators were modeled as ordered binary variables, and missing data were handled using pairwise present estimation implemented within lavaan. Comparative SEM evaluation incorporated: (i) comparative fit index (CFI), (ii) Tucker-Lewis index (TLI), (iii) root mean square error of approximation (RMSEA), (iv) standardized root mean square residual (SRMR), (v) standardized latent path coefficients, and (vi) residual covariance structure.

Primary downstream interpretation focused on relative stability and directional consistency of latent covariance structure across alternative psychosis embedding strategies rather than large individual effect magnitudes. This interpretation framework was selected because population-scale psychiatric genomic analyses are expected to demonstrate highly distributed and individually modest genetic effects across heterogeneous psychiatric phenotypes. Latent path coefficients and phenotype-level factor loadings were subsequently visualized using integrated SEM heatmaps summarizing standardized path estimates across M1-M3 frameworks.

**Leave-one-indicator-out and reduced-factor sensitivity analyses**

Additional SEM sensitivity analyses were performed to evaluate robustness of latent-factor organization and genetic path covariance structure across alternative phenotype compositions.

Leave-one-indicator-out (LOO) analyses sequentially removed individual phenotypic indicators from the bifactor structure to evaluate stability of latent factor covariance structure and standardized genetic path estimates following removal of individual phenotype domains.

Additional reduced-factor analyses excluded mood and anxiety phenotypes to evaluate whether latent psychiatric/developmental structure remained stable following removal of highly prevalent affective phenotypes that could potentially dominate broader psychiatric covariance architecture.

Reduced-factor analyses additionally re-evaluated one-factor, correlated two-factor, and bifactor latent structures following exclusion of mood and anxiety phenotypes.

#### Statistical visualization and figure generation

Figures were generated using Python version 3.10 and R version 4.3. Visualization workflows incorporated matplotlib, seaborn, pandas, numpy, ggplot2, and custom plotting utilities. Visualization approaches included chromosomal interval maps, phenotype-level dot plots, radar plots, heatmaps, latent-structure schematics, SEM path visualizations, and covariate perturbation sensitivity plots. All figures and derived outputs exported outside the Researcher Workbench environment adhered to *All of Us* Controlled Tier privacy and export requirements.

#### Data governance and ethics

All analyses were performed under approved National Institutes of Health (NIH) *All of Us* Research Program Controlled Tier data-use protocols within the secure Researcher Workbench environment. Access to Controlled Tier genomic and EHR-linked participant data required institutional agreements, researcher credentialing, and completion of required NIH and *All of Us* data-use training.

All downstream analyses adhered to *All of Us* participant privacy, genomic governance, and export-control policies. Only aggregate summary statistics, privacy-compliant derived outputs, and non-identifiable figures were exported outside the Researcher Workbench environment. No participant-level genomic identifiers, structural variant identifiers, direct participant identifiers, dates of birth, or row-level protected genomic information were exported outside the controlled cloud environment.

All exported figures, tables, and derived summary outputs were manually reviewed to ensure compliance with *All of Us* disclosure-control policies and Controlled Tier governance requirements.

#### ***All of Us*** privacy and participant protections

The *All of Us* Research Program incorporates extensive participant privacy and genomic governance protections designed to minimize risk of participant re-identification while preserving utility of large-scale genomic and longitudinal health datasets for biomedical research.

Controlled Tier genomic analyses were conducted exclusively within the secure cloud-native Researcher Workbench environment. Direct participant identifiers are not accessible within the Researcher Workbench environment, and genomic analyses are governed through tiered access controls, researcher authentication requirements, and export-review procedures.

Because structural variant analyses may theoretically increase re-identification risk through rare genomic events, all downstream recurrent ND-CNV analyses used participant-level aggregate carrier status definitions rather than export of row-level structural variant calls or participant-linked genomic intervals.

All analyses presented in this study reflect aggregate statistical summaries rather than participant-level genomic reporting.

#### Artificial intelligence and computational assistance disclosure

Artificial intelligence-assisted large language model (LLM) tools were used during portions of manuscript drafting, code refinement, figure legend drafting, and organizational editing workflows. AI-assisted tools were not used to generate primary scientific results, perform statistical analyses, modify underlying datasets, generate latent-variable model outputs, or autonomously interpret unreviewed participant-level genomic data.

All genomic analyses, statistical modeling procedures, structural equation modeling workflows, and downstream scientific interpretations were designed, executed, reviewed, and validated by the study authors within the *All of Us* Researcher Workbench environment.

All AI-assisted outputs incorporated into manuscript drafting or figure organization were manually reviewed, edited, validated against primary analytic outputs, and verified for scientific accuracy by the authors prior to inclusion in the final manuscript.

AI-assisted tools functioned exclusively as supportive drafting and organizational aids and did not replace direct investigator oversight, statistical validation, or scientific interpretation.

### II. Code Description for AoU ND-CNV, Schizophrenia PRS, and Logistic Regression Pipeline

#### Overview of the two-notebook workflow

This analysis was implemented as a two-notebook *All of Us* Researcher Workbench pipeline. Notebook 1 constructs the analytic cohort, derives electronic health record phenotypes, imports and harmonizes curated recurrent neurodevelopmental copy-number variant intervals, extracts observed structural variant calls from *All of Us* whole-genome sequencing MatrixTables, assigns participant-level ND-CNV exposures, and exports a minimal model-ready handoff table. Notebook 2 uses that handoff table to annotate recurrent ND-CNV calls with gene-set resources, calculate schizophrenia polygenic risk scores using AoUPRS, residualize and standardize the PRS, define psychosis-spectrum and RDoC-informed outcomes, run the primary and sensitivity logistic regression models, generate publication figures, and export AoU-safe result tables and SEM-ready files.

The logic of the pipeline is to separate high-cost genomic data processing from downstream association modeling. Notebook 1 performs Hail-based structural variant extraction and participant-level exposure construction. Notebook 2 works primarily from the compact exported participant-level table and the ro30-positive call-locus table, allowing downstream PRS calculation, annotation, modeling, visualization, and export to proceed without repeatedly scanning the full structural variant MatrixTables.

#### Notebook 1: Cohort construction, phenotype derivation, structural variant extraction, and participant-level ND-CNV exposure assignment

#### Environment setup, global parameters, and workflow configuration

Notebook 1 begins by initializing an AoU-native Hail genomics environment configured for Dataproc-based analysis. The recorded environment uses the *All of Us* Hail Genomics Analysis environment with 4 CPUs, 60 GB driver RAM, and two workers with 26 GB RAM and 150 GB disk each. The notebook defines workspace-specific cloud paths using the *All of Us* workspace bucket, including a project tag, checkpoint directory, and export directory. These cloud paths allow expensive intermediate Hail Tables to be checkpointed and reused.

The global parameters define the versioned project namespace, run date, curated interval assets, overlap rules, structural variant quality-control filters, cohort inclusion defaults, and minimum sample thresholds for downstream modeling. The primary interval set is `ND_RAW`, the primary overlap rule is `ro30`, and additional overlap rules include any-overlap, 50% and 70% reciprocal overlap, midpoint containment, and combined reciprocal-overlap/midpoint rules. Structural variant filtering is restricted to deletions and duplications, autosomal variants, variants at least 5 kb in size, and PASS-only calls. Cohort-level defaults require at least 365 days of observation time, and phenotype case definitions are designed around repeated clinical evidence, including at least two occurrences and distinct dates where available. The notebook also defines modeling feasibility thresholds of at least 20 exposed participants for locus-specific modeling and at least 10 cases per outcome.

This setup makes the pipeline reproducible by centralizing the values that control exposure definition, phenotype strictness, structural variant filtering, and file locations. It also ensures that each output is tied to a specific project tag and run version.

**Section A: Curated interval asset import and harmonization**

Section A imports the curated recurrent ND-CNV interval resources and matched-control interval resources. The primary ND-CNV interval files include raw GRCh38 recurrent ND-CNV loci without padding and a 100 kb padded version used for optional demonstration or sensitivity checks. The matched-control interval files include segmental-duplication and GC-matched control intervals corresponding to the raw ND-CNV loci, with a separate padded-control file available but not imported by default.

The code defines helper functions for importing BED-like interval files, harmonizing contig names, coercing start and end positions to numeric coordinates, calculating interval lengths, and assigning stable interval identifiers. It uses frozen GRCh38 contig lengths to prevent intervals from exceeding chromosome boundaries and to enforce valid genomic coordinate ranges. Each interval resource is converted into a Hail Table with standardized fields such as chromosome, start, end, interval ID, interval set, interval family, locus name, CNV direction when available, and target interval length.

The purpose of this section is to create a unified interval table containing all target recurrent ND-CNV loci and matched controls in a common schema. This harmonized interval table is checkpointed for reuse and forms the target interval payload used later for structural variant overlap assignment.

**Section B: Cohort definition, demographics, ancestry, relatedness, phenotype dictionary expansion, observation time, and healthcare utilization**

Section B constructs the canonical participant-level analytic cohort. The code first pulls person-level demographics from the All of Us OMOP `person` table, including participant identifiers, date of birth, sex-at-birth or gender concept identifiers where available, race and ethnicity concept identifiers, and demographic labels. Age variables are later derived relative to observation time or last observation.

The phenotype derivation begins with an uploaded ND-CNV psychiatric phenotype dictionary, `nd_cnv_phenotype_dictionary_20260406.csv`. This dictionary maps psychiatric and neurodevelopmental phenotypes to OMOP condition concepts and organizes them into phenotype names, labels, and families. The code cleans and validates the dictionary, then uses BigQuery against the OMOP concept-ancestor tables to expand each seed concept to descendant condition concepts. This descendant expansion is crucial because EHR conditions may be recorded under more specific descendant concepts rather than the exact seed concept listed in the dictionary. After expansion, the code pulls long-format `condition_occurrence` records for all dictionary-derived descendant concepts and merges those condition rows back to the phenotype metadata.

The long-format condition records are collapsed into participant-level phenotype flags. For each participant and phenotype, the code computes whether the phenotype was ever observed, the number of condition records, the number of distinct condition start dates, the number of distinct OMOP concepts, and the first observed date. These repeated-code summaries are retained because they support later sensitivity analyses using stricter phenotype definitions. The final phenotype table is wide-format, with one row per participant and binary columns for each phenotype name.

The notebook also pulls observation-period summaries and healthcare-utilization summaries. Observation time is used to quantify longitudinal EHR capture, while healthcare utilization is summarized through counts of clinical visits and log-transformed visit counts. These covariates are important because participants with more healthcare contact have more opportunities to receive diagnostic codes, creating ascertainment differences that could confound EHR-based association analyses.

Genomic QC data are then imported from *All of Us* workspace resources, including genomic metrics, flagged sample lists, relatedness exclusions, and ancestry predictions with genetic principal components. The code applies sex-concordance and sample-level QC filters, removes flagged or related samples as appropriate, joins ancestry and PC data, and assembles a canonical participant table. Before converting this table to Hail, datetime columns are converted to stable ISO-formatted strings to avoid Hail import errors related to pandas timestamp parsing. The resulting Hail Table, `ht_samples`, is keyed by `person_id` and checkpointed as the cohort anchor for structural variant processing.

**Section C: Kernel-safe extraction of observed structural variant calls from ND-CNV chromosome MatrixTables**

Section C extracts compact structural variant call records from per-chromosome *All of Us* ND-CNV MatrixTables. The notebook first inspects a representative MatrixTable schema to identify row fields, column fields, and entry fields. The row fields include genomic locus, alleles, filters, nested INFO fields, structural variant type, and deletion/duplication indicators. The entry fields include genotype and copy-number-related fields such as `GT`, `CN`, `CNQ`, `GQ`, read-depth genotype fields, and split-read or paired-end support fields.

The code defines per-chromosome ND-CNV MatrixTable paths and builds a per-chromosome curated interval payload. It then iterates over relevant autosomes and extracts only compact observed call information needed for overlap testing. Filtering retains autosomal PASS-only deletions and duplications meeting the minimum size threshold. The extracted fields include participant ID, chromosome, start, end, SV type, observed call length, call-level identifiers, and available genotype or copy-number support fields.

The logic of this section is to avoid materializing unnecessary full MatrixTable data. Instead, each chromosome-level MatrixTable is reduced to a compact Hail Table of observed SV calls, checkpointed by chromosome, and then unioned across chromosomes. A stable `observed_call_id` is assigned so that each participant-level observed call can be tracked through overlap, collapse, and audit steps. The final output is a long-format table of observed SV calls, `ht_sv_calls_long_with_ids`, that is suitable for interval overlap testing.

**Section D: Structural variant by curated interval overlap assignment and collapse**

Section D intersects observed structural variant calls with the curated recurrent ND-CNV and matched-control intervals. Before overlap assignment, the observed-call table is cleaned and restricted to the main analysis interval families. The code computes interval overlap metrics including base-pair overlap length, fraction of the observed call overlapped, fraction of the target interval overlapped, reciprocal overlap status, and midpoint containment. It then assigns boolean flags for each pre-specified overlap rule: any overlap, ro30, ro50, ro70, midpoint, ro30 plus midpoint, and ro50 plus midpoint.

The primary exposure definition is ro30, meaning that a participant is considered exposed at a curated locus when an observed deletion or duplication overlaps the target interval by the required reciprocal-overlap threshold. The notebook also retains the other overlap rules for audit and sensitivity documentation, but the ro30 rule is used as the primary downstream definition.

The code creates a dedicated ro30-positive ND call-locus table for Notebook 2, `ht_nd_call_locus_ro30_for_n2`. This table has one row per observed CNV call by curated ND locus under the primary overlap rule and retains genomic coordinates required for gene annotation. This is the key coordinate-level handoff from Notebook 1 to Notebook 2.

The notebook then collapses overlap candidates to participant-level evidence. ND-CNV overlaps are collapsed across observed calls and intervals, while matched-control overlaps are restricted immediately to ro30-positive rows to reduce computational burden. For the matched-control branch, the code uses sharding and two-stage aggregation to avoid memory pressure from the larger control interval space. ND and control collapsed tables are harmonized to a shared schema, unioned, and annotated with `passed_rules`, an array of overlap rules satisfied by each participant-interval observation.

Diagnostic summaries are generated to audit rule-combination frequencies, participant exposure counts by rule, deletion/duplication mixing, and locus-specific exposed participant counts under ro30. Loci with at least 20 exposed participants are identified as potentially feasible for locus-specific modeling, although the primary manuscript analyses rely on aggregate recurrent ND-CNV carrier status.

**Section E: Participant-level exposure table construction**

Section E converts interval-level overlap evidence into participant-level exposure variables. The code starts from the lean overlap table produced in Section D and annotates each row with rule-specific indicators. It then deduplicates to ensure one row per participant, interval set, and interval ID before aggregating to participant-level features.

The participant-level exposure table includes binary indicators for any ND-CNV exposure, deletion exposure, duplication exposure, matched-control exposure, and rule-specific exposure status. It also includes counts of exposed intervals and exposed loci where appropriate. Missing exposure values are filled as zero or false for unexposed participants. The exposure table is left-joined onto the full canonical cohort so that every analytic participant remains represented, not only participants with observed CNVs.

This section produces the key participant-level analytic table, `ht_analysis_cohort_with_exposure`, which is then exported to pandas as `df_analysis`. The code performs defensive type normalization after Hail-to-pandas conversion, ensuring that exposure flags are clean binary variables and that count variables are numeric. It also performs assertions to confirm one row per participant, nonnegative counts, no missing exposure indicators, and expected behavior of control exposures.

**Section F/G setup: Phenotype and covariate preparation for downstream modeling**

After exposure construction, Notebook 1 prepares dictionary-driven phenotype helpers and covariate lists for downstream analysis. It identifies which phenotype flags survived into `df_analysis`, coerces phenotype flags to binary integers, and preserves repeated-code count, date, and concept summaries. It also builds a family-label map so that downstream models and figures can group outcomes by phenotype family.

The initial covariate setup includes demographic, ancestry, and healthcare-utilization variables. Although Notebook 2 ultimately defines the final logistic regression covariate sets, Notebook 1 prepares the columns needed for those models, including age, sex-at-birth concept information, observation time, visit counts, ancestry labels, and genetic principal components.

**Minimal transfer export from Notebook 1 to Notebook 2**

The final Notebook 1 section builds a compact handoff package for downstream PRS and logistic modeling. The central export is `df_model_base.csv`, which includes one row per analytic participant, demographic and ancestry covariates, participant-level ND-CNV exposure variables, phenotype flags, repeated-code phenotype summaries, healthcare-utilization variables, and observation-time variables. The export intentionally preserves `sex_at_birth_concept_id` so that Notebook 2 can recode sex consistently for regression and SEM rather than treating raw OMOP concept IDs as quantitative variables.

Notebook 1 also exports `df_analysis.csv`, `phenotype_dict_df.csv`, `phenotype_screen_df.csv`, `family_label_map_df.csv`, and `notebook1_analytic_person_ids.tsv`. The person ID file is formatted for PLINK compatibility with FID set to 0 and IID set to `person_id`, reflecting the expected *All of Us* sample ID convention. The notebook records the checkpoint paths for `ht_nd_call_locus_ro30_for_n2`, `ht_overlap_for_exposure_with_rule_flags`, `ht_participant_exposure`, and `ht_samples`, allowing Notebook 2 to access both compact participant-level and coordinate-level outputs without rerunning the full structural variant extraction pipeline.

#### Notebook 2: Functional annotation, AoUPRS scoring, logistic regression, figures, and AoU-safe exports

##### Environment setup and Notebook 1 handoff import

Notebook 2 begins by initializing local and cloud working directories for PRS and modeling outputs. It then loads the minimal Notebook 1 handoff table, typically `df_model_base.csv`, into pandas as `df_analysis`. The code verifies that required participant-level exposure, covariate, and phenotype fields are present.

The notebook also rebuilds RDoC primary flags if they are absent. These RDoC flags summarize phenotype dictionary-derived outcomes into broader Research Domain Criteria-informed domains. The rebuilt domain flags allow the same participant-level table to support both phenotype-level logistic regression and dimensional RDoC-informed models.

##### Section B: External asset setup and preflight

Notebook 2 requires several external resources: the schizophrenia PRS scoring file, GENCODE gene annotations, gnomAD constraint metrics, SFARI gene annotations, and PGC schizophrenia genes. The code defines user-editable local filenames or existing Google Cloud Storage paths for these files, then resolves each asset to a canonical workspace-bucket path. If a file is present locally but not yet in the bucket, the helper function uploads it using `gsutil`. If an input is already a `gs://` path, the code verifies that it exists.

The required external assets are `PGS002785_hmPOS_GRCh38.txt.gz` for schizophrenia PRS weights, `gencode.v38.annotation.gff3.gz` for gene coordinates, `gnomad.v4.1.1.constraint_metrics.tsv.bgz` for loss-of-function constraint, `SFARI-Gene_genes_03-22-2026release_04-22-2026export.csv` for autism and neurodevelopmental gene annotations, and `NIHMS59304-supplement-Supplementary_Table_4.xlsx` for PGC 2014 schizophrenia-prioritized loci. The preflight step confirms that all required files are accessible before the notebook proceeds.

##### Section C: Gene intervals and gene-set resources

Section C loads GENCODE v38 gene intervals from the GFF3 file and parses gene-level records into a table with chromosome, start, end, gene symbol, gene ID, and other annotation fields. These gene intervals provide the coordinate framework for intersecting recurrent ND-CNV calls with genes.

The notebook then loads gene-set resources. Loss-of-function intolerant genes are derived from gnomAD v4.1.1 constraint metrics using LOEUF-based filtering on MANE Select or canonical transcripts, with LOEUF < 0.35 used as the high-constraint threshold. SFARI genes are parsed into primary autism-associated genes, typically SFARI scores 1–2, and a broader neurodevelopmental sensitivity set, typically SFARI scores 1–3. PGC schizophrenia genes are read from the 2014 schizophrenia loci workbook without relying on openpyxl, allowing the code to extract schizophrenia-prioritized genes from the supplementary table.

This section creates gene-set membership objects that are later used to annotate ND-CNV carriers according to whether their recurrent CNV intervals overlap loss-of-function intolerant genes, SFARI genes, broad neurodevelopmental genes, or schizophrenia-prioritized loci.

**Section D: Annotation of ro30-positive ND-CNV calls with overlapping genes**

Section D loads the ro30-positive ND call-locus Hail Table generated by Notebook 1. This table contains one row per observed recurrent ND-CNV call by curated locus and retains the genomic coordinates required for gene overlap.

The code intersects these call-locus intervals with GENCODE gene intervals and annotates each overlapping gene with membership in the loaded gene sets. Each call-gene row is labeled according to whether the gene belongs to the loss-of-function intolerant set, SFARI primary set, SFARI broad neurodevelopmental set, PGC schizophrenia set, or related schizophrenia coding/eQTL-linked categories when available.

The call-gene annotations are collapsed to participant-level gene-set flags. For each participant, the notebook records whether any recurrent ND-CNV overlaps each gene-set category. These annotation flags are merged back into `df_analysis`, and noncarriers are assigned false/zero values. The resulting participant-level annotation variables support Figure 1-style summaries and supplement-ready gene annotation exports.

**Section E: AoUPRS-based schizophrenia PRS scoring**

Section E calculates schizophrenia polygenic risk scores using AoUPRS. AoUPRS is installed and configured inside the *All of Us* workspace environment. The scoring file used in this notebook is PGS002785 in GRCh38/hmPOS format. The code reloads the scoring file and creates a reduced AoUPRS-compatible weight table using the top 100 variants per chromosome. This reduction step makes the analysis computationally lightweight and tractable within the workspace while preserving a schizophrenia PRS signal suitable for manuscript-scale demonstration and modeling.

The reduced PRS weight table is written locally and uploaded to the workspace bucket. The notebook then loads the *All of Us* v8 Variant Dataset (VDS) and restricts the scoring operation to the analytic participants from Notebook 1. The analytic sample list is also written to the bucket for provenance and reuse.

AoUPRS is then run in VDS mode to calculate participant-level schizophrenia PRS. In *All of Us* v8, the WGS data are represented in a sparse VDS format rather than a dense MatrixTable, and AoUPRS is designed to operate efficiently on this representation. AoUPRS reads the reduced scoring file, matches scoring variants to the *All of Us* WGS VDS, uses available genotype representation in the VDS to compute weighted allele dosages, and returns participant-level PRS values. The raw output is normalized into `df_prs`, containing `person_id`, `prs_scz_raw`, and the number of variants contributing to the score, `N_variants`. The raw PRS output is saved internally as `df_prs_scz_raw.csv`.

A PRS QC summary is generated to document the number of scored participants, score distribution, and variant count distribution. These QC outputs are intended for internal audit and supplement-level reporting rather than row-level public sharing.

**Section F: Merge PRS, residualize, and standardize**

Section F merges `df_prs` into the participant-level analytic dataset. The raw schizophrenia PRS is first summarized within the model-ready cohort. The notebook then residualizes PRS to reduce confounding by demographic and population-structure covariates. Numeric covariates are coerced defensively, and available genetic principal components are detected automatically.

The primary residualized PRS variable is `prs_scz_resid_z`, a standardized residual from a model adjusting the raw PRS for covariates such as age, sex, ancestry, and principal components depending on column availability. The notebook also creates `prs_scz_z_within_ancestry`, a within-ancestry standardized PRS. This within-ancestry score is used as the primary SEM-aligned PRS when available and is also aliased as `prs_scz_sem_z` for later lavaan export. The purpose of these transformations is to put the PRS on a comparable standard-deviation scale while reducing confounding due to ancestry-related score distribution differences.

**Section G: Outcome schema and joint logistic regression models**

Section G defines the final outcome schema for schizophrenia, psychosis-spectrum phenotypes, broader psychiatric phenotypes, and RDoC-informed domains. The notebook explicitly avoids defining broad psychosis from overly broad dictionary groupings that may contain nonspecific neurologic or sensorimotor phenotypes such as numbness. Instead, it constructs psychosis phenotypes from named diagnosis and symptom columns.

The strictest schizophrenia phenotype, `schizophrenia_specific`, is defined using schizophrenia and paranoid schizophrenia flags. A broader `schizophrenia_spectrum` phenotype adds schizoaffective disorder. A broader psychotic-disorder phenotype includes schizophrenia, paranoid schizophrenia, schizoaffective disorder, psychotic disorder, and reactive psychosis. The primary `psychosis_broad` phenotype combines those disorder-level psychosis variables with symptom-level psychosis or thought-disorder features, including hallucinations, delusions, and disturbance of thinking. Motor or sensorimotor variables are explicitly excluded from this psychosis definition because they may reflect medication effects, nonspecific neurologic symptoms, or unrelated coding patterns.

The logistic regression covariate structure is then defined. Sex at birth is recoded into a centered binary covariate rather than raw OMOP concept IDs. The primary covariate set includes age at last observation, centered binary sex, log-transformed unique visit count, total observation time, and ancestry principal components PC1–PC5. Two utilization sensitivity covariate sets are also created. One uses healthcare utilization residualized on age, observation time, sex, and PCs. The other uses utilization decile calculated within age bands and then standardized. These covariate sets allow the analysis to test whether associations are robust to different approaches for controlling EHR ascertainment.

The core logistic regression helper, `run_logit_safe`, creates a complete-case modeling dataset for each outcome, predictor set, and covariate set. It coerces all columns to numeric, drops missing or infinite values, checks that the model has at least 100 observations and at least 10 cases, removes constant predictors, fits a logistic regression model, and returns odds ratios, confidence intervals, p-values, sample size, case count, model status, and diagnostic messages. The helper is designed to fail safely and return an interpretable skipped-model row rather than crashing when outcomes are too rare or predictors are noninformative.

The primary model specification includes `nd_any`, labeled as any recurrent ND-CNV. Direction-specific exploratory model specifications include `nd_del_any` and `nd_dup_any`. For each outcome and exposure, the code creates an interaction term between the exposure and the primary PRS column. The base predictor set for joint models is therefore the ND-CNV exposure, schizophrenia PRS, and the PRS-by-ND-CNV interaction term, adjusted for the selected covariates.

The notebook runs several model families. The primary models use any ND-CNV carrier status and the raw-utilization covariate set. Direction-specific exploratory models repeat the framework using deletion and duplication carrier status. Utilization sensitivity models rerun any-ND-CNV analyses using residualized utilization and age-binned utilization decile covariates. Internalizing-adjusted psychosis sensitivity models add internalizing phenotypes as additional covariates when evaluating psychosis outcomes. Repeated-code sensitivity models construct stricter outcomes using the repeated-code count/date/concept fields exported from Notebook 1, when those columns are available.

After all model families are run, the code combines the outputs into `df_results_core`, applies false-discovery-rate correction within model families, and exports several model-specific result tables. Core exports include `df_results_core_prs_cnv_models.csv`, `df_results_core_prs_cnv_sensitivity_models.csv`, `df_results_core_primary.csv`, `df_results_core_directional.csv`, `df_results_core_utilization_sensitivity.csv`, `df_results_core_internalizing_adjusted.csv`, and `df_results_core_repeated_code_sensitivity.csv`.

**RDoC-informed domain modification models**

After phenotype-level models, the notebook runs exploratory RDoC primary-domain models. These models use the same PRS, ND-CNV exposure, interaction, and covariate logic but apply it to RDoC-informed binary domain flags rather than individual diagnoses. The output is `df_results_rdoc`, with domain-level outcomes, terms, odds ratios, confidence intervals, p-values, sample sizes, case counts, model family labels, and FDR-adjusted q-values.

Direction-specific RDoC models are also exported as `df_results_rdoc_directional.csv`. These domain-level models support the dimensional analyses in the manuscript and allow comparison of common polygenic liability, any ND-CNV carrier status, deletion carrier status, duplication carrier status, and PRS-by-CNV interaction effects across broader neurobehavioral systems.

**Figure generation**

Notebook 2 generates both intermediate and publication-quality figures. The PRS distribution figure visualizes the schizophrenia PRS distribution and overlays ND-CNV carrier status to evaluate whether carrier and noncarrier PRS distributions are broadly comparable after harmonization. The primary regression forest plot visualizes odds ratios and confidence intervals for key PRS, ND-CNV, and interaction terms across psychosis and related phenotypes. The RDoC heatmap visualizes dimensional PRS and CNV modification signals across primary RDoC domains.

The publication figure section defines consistent plotting helpers and then writes figure-ready PNGs to a `figures_publication` directory. Outputs include `figure_prs_distribution_ndcnv_overlay.png`, `figure_ndcnv_functional_annotation_profile.png`, `figure_primary_regression_forest_prs_ndcnv.png`, `figure_rdoc_prs_modification_heatmap.png`, and `figure_main_composite_ndcnv_prs_primary_analysis.png`. The composite figure combines the PRS distribution, functional annotation profile, primary regression forest plot, and RDoC heatmap into a main manuscript figure.

**Section H: AoU-safe result tables and supplement-ready exports**

Section H creates AoU-safe export tables designed to summarize the analysis without releasing row-level participant identifiers or restricted individual-level genomic data. The CNV ascertainment and QC supplement table, `df_cnv_ascertainment_qc_supplement.csv`, summarizes recurrent ND-CNV exposure architecture by locus and CNV type using the ro30 call-locus table when available. If locus-level fields are unavailable, it falls back to participant-level aggregate exposure counts.

The carrier prevalence audit table, `df_cnv_carrier_prevalence_audit.csv`, summarizes ND-CNV, deletion, and duplication carrier frequencies across demographic, ancestry, age, sex, and utilization strata. This table is intended to document possible ascertainment or demographic differences in recurrent ND-CNV carrier prevalence.

The effect landscape table, `df_effect_landscape_psychosis_rdoc_for_figures.csv`, combines PRS, any ND-CNV, deletion, duplication, and PRS-by-CNV interaction effects across psychosis and RDoC primary outcomes. This table supports manuscript figures comparing schizophrenia polygenic liability and structural genomic carrir effects within a single harmonized framework.

The gene-set resource summary table, `df_gene_set_resource_summary.csv`, reports the sizes and provenance of external annotation resources used for functional interpretation. The participant-level gene annotation summary, `df_ndcnv_gene_annotation_carrier_summary.csv`, summarizes how many ND-CNV carriers overlap each gene-set category. The locus-level gene annotation summary, `df_ndcnv_gene_annotation_locus_summary.csv`, summarizes gene overlaps at the locus or interval level where available.

**SEM-ready export and lavaan model syntax generation**

Although the R SEM modeling is run outside these logistic-regression notebooks, Notebook 2 creates the SEM-ready input dataset and starter lavaan model syntax. The SEM export is designed to be ID-free and lavaan-compatible. The code selects primary psychiatric and developmental indicators, the primary PRS scale, ND-CNV exposure variables, interaction terms, covariates, and sex recoded as a centered binary variable. Binary indicators are coerced to 0/1 integers, exposure variables are coerced to 0/1 integers, and numeric covariates are forced to numeric type. The final SEM dataset intentionally excludes person IDs or sample identifiers and is exported as `df_sem_lavaan_internal_no_ids.csv`.

The notebook writes an R syntax file, `lavaan_general_psych_models.R`, containing starter lavaan specifications for three models. Model 1 defines a general psychiatric/developmental liability factor excluding psychosis. Model 2 models psychosis downstream of the non-psychosis latent liability factor. Model 3 includes psychosis as a factor-loading indicator. The syntax also includes a shared covariate block and notes that direction-specific sensitivity models can be created by replacing `nd_any` and `prs_x_nd_any` with deletion- or duplication-specific exposure and interaction terms.

Additional SEM-related summary exports include `df_sem_indicator_counts.csv` and `df_sem_exposure_counts.csv`. These tables document the count and prevalence of SEM indicators and exposure variables in the SEM dataset.

**Final model and PRS summary exports**

The notebook concludes by exporting all primary logistic regression, RDoC, sensitivity, PRS, and model-sample summary tables to the local exports directory and then copying them to the workspace bucket. The primary exported model outputs are `df_results_core_prs_cnv_models.csv`, `df_results_rdoc_prs_cnv_models.csv`, `df_results_core_prs_cnv_sensitivity_models.csv`, `df_results_core_primary.csv`, `df_results_core_directional.csv`, `df_results_core_utilization_sensitivity.csv`, `df_results_core_internalizing_adjusted.csv`, `df_results_core_repeated_code_sensitivity.csv`, and `df_results_rdoc_directional.csv`.

The notebook also exports `df_logistic_outcome_counts.csv`, which summarizes outcome case counts for the logistic regression phenotypes; `df_model_sample_counts.csv`, which summarizes available model sample sizes; and `df_prs_scz_distribution_summary.csv`, which summarizes the schizophrenia PRS distribution without exporting row-level PRS values. These outputs make the analysis auditable while remaining consistent with *All of Us* data governance expectations.

**Reprodicibility logic of the full pipeline**

The complete analysis comprises two primary All of Us Researcher Workbench notebooks, a standalone broad FINEMAP–ND-CNV genomic annotation package, a local reduced-PRS validation workflow, and a local PRS-decile validation package. Notebook 1 begins from versioned curated interval files, the phenotype dictionary, *All of Us* OMOP tables, genomic QC files, ancestry predictions, relatedness exclusions, and per-chromosome structural variant MatrixTables. It produces compact participant-level exposure and phenotype tables, plus a coordinate-level ro30 ND-CNV call-locus table for annotation. Notebook 2 begins from those handoff objects, adds external annotation resources and schizophrenia PRS weights, calculates PRS using AoUPRS, residualizes and standardizes PRS, defines final outcomes and covariates, runs primary and sensitivity logistic regression models, creates figures, and exports supplement-ready and SEM-ready datasets.

The central analytic design choices are defined as follows. Recurrent ND-CNV exposure is defined primarily using participant-level interval-collapsed ro30 overlap rather than raw row-level SV call carrier status. Phenotypes are dictionary-driven and descendant-expanded through OMOP vocabulary tables. Healthcare-utilization and observation-time covariates are included to reduce EHR ascertainment bias. Genetic ancestry is addressed using principal components and within-ancestry PRS standardization. PRS and CNV effects are modeled jointly with an interaction term, allowing the pipeline to test common polygenic effects, rare structural variant effects, and PRS-by-CNV interaction effects within the same regression framework. Sensitivity analyses evaluate robustness to direction-specific CNV carrier status, utilization adjustment strategy, internalizing adjustment, and stricter repeated-code phenotype definitions.

Taken together, the two notebooks implement an end-to-end *All of Us* workflow for studying the relationship between recurrent neurodevelopmental CNVs, schizophrenia polygenic risk, and EHR-derived psychiatric phenotypes. A researcher reproducing this pipeline would need the same curated recurrent ND-CNV interval files, matched-control interval files, phenotype dictionary, *All of Us* v8 controlled-tier OMOP and WGS resources, genomic QC and ancestry resources, external gene-set annotation files, and PGS002785 schizophrenia scoring file. They would then run Notebook 1 to generate the participant-level analytic handoff and ro30 call-locus table, followed by Notebook 2 to annotate ND-CNV calls, calculate schizophrenia PRS with AoUPRS, construct model-ready predictors and outcomes, run logistic regression and sensitivity analyses, and export manuscript-ready result tables and figures.

### III. External notebooks and locally executed analysis packages

Several downstream validation and genomic benchmarking analyses were conducted outside the All of Us Researcher Workbench using standalone Python notebooks or reproducible local analysis packages. These workflows did not require access to identifiable participant-level data or raw protected genomic calls. The external workflows used either publicly available genomic annotation resources, curated interval files generated during the primary analysis, or ID-free model-ready summary datasets exported in accordance with All of Us Controlled Tier privacy requirements. They were maintained separately from the two-notebook Workbench pipeline to permit independent rerunning of figure-generation, validation, and annotation procedures without rescanning the All of Us WGS structural-variant resources or recalculating participant-level PRS.

#### Broad FINEMAP-ND-CNV overlap analysis

Figure 1B was generated using a standalone local Python workflow rather than Notebook 2 of the All of Us Researcher Workbench pipeline. The purpose of this analysis was to benchmark the curated recurrent ND-CNV interval framework against the broader schizophrenia GWAS discovery landscape using the FINEMAP-derived gene set reported by Trubetskoy et al.

The workflow used three primary inputs. First, the curated recurrent ND-CNV BED file contained 97 deletion- and duplication-labeled interval rows harmonized to GRCh38 coordinates. After exclusion of the chromosome X interval, 96 autosomal rows remained. Because reciprocal deletion and duplication entries commonly shared identical genomic boundaries, rows were collapsed by chromosome, start coordinate, and end coordinate to yield 51 unique autosomal genomic intervals. The deletion- and duplication-specific locus labels associated with each unique interval were retained in the summary outputs.

Second, the schizophrenia annotation input consisted of the reconstructed broad FINEMAP set of 435 protein-coding genes. This set was derived from the Trubetskoy et al. Supplementary Table 11b 95% credible-set annotations by retaining unique protein-coding Ensembl genes. The reconstruction reproduced the published total of 435 protein-coding FINEMAP genes and was used as a broad discovery-oriented resource distinct from the more stringently prioritized schizophrenia gene resources used in the Workbench-based Figure 1C and Figure 1D analyses.

Third, GENCODE release 38 gene annotations were used to obtain GRCh38 gene-body coordinates. Only gene records on chromosomes 1–22 were retained. GFF3 coordinates, originally represented as 1-based inclusive intervals, were converted to 0-based half-open coordinates to match the BED interval convention. Version suffixes were removed from Ensembl identifiers before matching. Broad FINEMAP genes were matched to GENCODE primarily by stable Ensembl gene identifier, with gene-symbol matching used as a secondary fallback. Of the 435 broad FINEMAP genes, 429 were successfully matched to GENCODE v38; six identifiers were not matched and were retained in a dedicated audit table.

Direct gene-body overlap was defined when the GENCODE gene interval and curated recurrent ND-CNV interval shared at least one base pair. No additional upstream, downstream, or interval padding was applied. The workflow iterated over autosomes, identified all broad FINEMAP genes overlapping each unique recurrent ND-CNV interval, and removed duplicate interval–gene combinations. Overlap length in base pairs was also calculated for each interval–gene pair.

The resulting interval–gene table was aggregated in two directions. At the locus level, the workflow calculated the number of unique broad FINEMAP genes intersecting each recurrent ND-CNV interval, the corresponding gene symbols, the interval length in base pairs, an indicator for whether the interval contained at least one FINEMAP gene, and FINEMAP gene density normalized per megabase of interval length. At the gene level, the workflow calculated the number and identities of recurrent ND-CNV intervals intersecting each broad FINEMAP gene.

Across the 435-gene broad FINEMAP set, 28 genes intersected at least one curated autosomal recurrent ND-CNV interval. These 28 genes represented 6.4% of the complete broad protein-coding FINEMAP set. Conversely, 13 of the 51 unique autosomal recurrent ND-CNV intervals contained at least one broad FINEMAP gene, corresponding to 25.5% of the unique interval framework.

The primary Figure 1B visualization was generated from the complete 51-interval locus-level table. Each bubble represents one unique autosomal recurrent ND-CNV interval. The x-axis represents the number of overlapping broad FINEMAP genes, and the y-axis represents broad FINEMAP gene density, calculated as the number of overlapping genes divided by interval length in megabases. Both bubble area and continuous color encode interval length in megabases. All 51 intervals are plotted, including intervals with zero FINEMAP-gene overlap, while labels are restricted to the 13 intervals containing one or more overlapping genes. Automated label adjustment was used to reduce text collisions and retain labels within the plotting area.

The standalone package generated the following primary tables:

- `figure1b_overlap_global_summary.csv`, containing the total broad FINEMAP gene count, number mapped to GENCODE v38, number and percentage intersecting recurrent ND-CNV intervals, total number of unique autosomal ND-CNV intervals, and number and percentage containing at least one FINEMAP gene.
- `exhaustive_ndcnv_broad_finemap_gene_intersections.csv`, containing one row per unique recurrent ND-CNV interval–FINEMAP gene overlap, including interval coordinates, retained locus labels, Ensembl gene ID, gene symbol, GENCODE gene coordinates, gene type, and overlap length.
- `ndcnv_locus_level_broad_finemap_summary.csv`, containing all 51 unique autosomal recurrent ND-CNV intervals, interval length, number and list of overlapping FINEMAP genes, binary overlap status, and gene density per megabase.

- `broad_finemap_gene_level_ndcnv_overlap_summary.csv`, containing the 28 intersecting broad FINEMAP genes and the number and identities of recurrent ND-CNV intervals intersecting each gene.
- `broad_finemap_genes_unmatched_to_gencode_v38.csv`, containing the six broad FINEMAP Ensembl identifiers that could not be mapped to GENCODE release 38.

The package additionally produced exploratory figures summarizing raw gene counts by locus, the distribution of gene counts across intervals, genomic distribution across autosomes, and locus-level gene density. The final manuscript-facing Figure 1B bubble plot combines the locus-level gene-count and interval-normalized density information while retaining interval length as an additional visual variable.

The complete workflow is implemented in `run_figure1b_overlap.py` and wrapped by `figure1b_broad_finemap_ndcnv_overlap.ipynb`. The notebook imports and executes the standalone script, displays the global summary, and presents the highest-overlap recurrent ND-CNV intervals for audit. The underlying script contains the complete coordinate conversion, gene matching, interval intersection, aggregation, export, and figure-generation procedures.

**Local validation of the reduced schizophrenia PRS index**

Because the schizophrenia PRS used in the primary analysis was implemented as a computationally reduced index containing the highest-weighted variants from each autosome, a separate local validation workflow was used to document the construction and composition of the reduced scoring set. This workflow was designed to evaluate whether the reduction procedure preserved genome-wide representation, balanced positive and negative effect weights, and selected variants from the high-weight tail of the original PGS Catalog score without allowing a small number of variants to dominate the reduced index.

The analysis began with the harmonized PGS Catalog scoring file for PGS002785. Metadata comment lines were parsed and retained separately, while the tab-delimited variant-level scoring table was loaded into pandas. Required fields included rsID, effect allele, effect weight, harmonized chromosome, harmonized genomic position, and inferred alternate allele. Chromosome labels were normalized, and analyses were restricted to chromosomes 1–22.

The validation workflow applied the same reduction criteria used for the Workbench PRS implementation. Variants were required to have valid GRCh38 harmonized positions, numeric effect weights, and unambiguous single-nucleotide A/C/G/T effect and alternate alleles. Variants meeting these criteria were ranked within each autosome according to the absolute magnitude of the reported effect weight. The top 100 variants from each autosome were retained, yielding a fixed 2,200-variant reduced PRS index.

The original PGS002785 scoring file contained 964,422 rows, of which 964,355 mapped to autosomes. After applying the harmonized-position and conservative biallelic allele filters, 761,361 usable autosomal variants remained. The final 2,200-variant index therefore represented 0.23% of the complete scoring file and contained exactly 100 variants from each of the 22 autosomes.

Several complementary quality-control analyses were conducted. The absolute-weight distribution of the 2,200 retained variants was compared with that of the full usable autosomal background. The reduced set exhibited a higher mean absolute effect weight than the usable background, as expected from the selection procedure. Specifically, mean absolute weight was approximately 0.000641 in the reduced set compared with 0.000100 in the usable background, while median absolute weights were approximately 0.000497 and 0.000082, respectively.

Chromosome-level summaries documented the number of retained variants, signed and absolute-weight distributions, genomic coordinate range, and minimum and maximum retained effect weight for each autosome. The fixed top-100-per-autosome strategy ensured complete autosomal representation rather than permitting chromosomes with larger numbers of scoring variants to dominate the reduced score.

Signed effect-weight balance was evaluated in both the complete usable background and reduced index. The reduced index contained 50.7% positive and 49.3% negative weights, compared with 50.1% positive and 49.9% negative weights in the usable background. This indicated that selecting variants by absolute effect magnitude did not produce substantial directional imbalance.

Weight-concentration analyses quantified the proportion of the total absolute weight contributed by the largest retained variants. These analyses were used to determine whether the reduced score was dominated by a small number of unusually large weights or whether its aggregate weight remained distributed across the selected variants. The genomic distribution panel additionally plotted retained absolute weights across cumulative autosomal position to verify that selected variants were distributed throughout the genome rather than confined to a small number of genomic regions.

Where ID-free participant-level summary data were available, the notebook also evaluated the distributions of the standardized PRS variables used in downstream analyses and correlations between those PRS variables and ancestry principal components. The primary SEM-aligned score, `prs_scz_sem_z`, included 71,992 participants, had a mean near zero and standard deviation near one, and ranged from approximately −4.61 to 3.97. The workflow similarly summarized `prs_scz_resid_z` and `prs_scz_z_within_ancestry`. Optional PRS–principal-component correlation tables were generated to document residual relationships between the harmonized scores and PC1–PC5.

The primary source and audit tables include:

- PGS002785_reduced_top100_per_autosome_GRCh38.csv and .tsv, containing the 2,200 retained variants, effect alleles, signed effect weights, harmonized coordinates, absolute weights, and within-chromosome weight ranks.
- reduced_prs_variant_qc_summary.csv, containing scoring-file metadata and counts at each filtering and reduction stage.
- reduced_prs_per_chromosome_summary.csv, containing retained-variant counts, signed and absolute-weight summaries, and genomic coordinate ranges for each autosome.
- reduced_prs_top50_variants_by_abs_weight.csv, containing the 50 largest absolute-weight variants retained in the reduced score.
- background_vs_reduced_summary.csv, comparing variant count, mean, median, upper-tail, and maximum absolute weights, along with positive and negative weight proportions, between the usable background and reduced index.
- panel_A_reduction_summary.csv, documenting the progression from the complete PGS scoring file to the 2,200-variant index.
- panel_C_chromosome_representation.csv, documenting usable background and retained scoring variants by chromosome.
- panel_D_signed_weight_balance.csv, summarizing positive and negative effect-weight percentages.
- panel_E_weight_concentration.csv, summarizing the contribution of the largest retained variants to total absolute score weight.
- PGS002785_reduced_top100_per_autosome_source_data.csv, containing the complete plotting and QC variables for all retained variants.
- optional_prs_distribution_summary_copy.csv and optional_prs_pc_correlations.csv, containing participant-level distribution summaries and PRS–PC correlations when those ID-free inputs were available.

The package contains two related notebooks. PRS_reduced_score_validation_local_notebook.ipynb performs the complete reconstruction, filtering, variant-level quality control, export, and optional participant-level PRS checks. PRS_reduced_PRS_validation_figure_notebook.ipynb generates the manuscript-facing validation panels and corresponding source tables.

The final multi-panel validation figure includes: the reduction funnel from the complete PGS file to the 2,200-variant score; the distribution of absolute weights in the usable background and reduced index; autosomal representation; signed weight balance; absolute-weight concentration; and genomic distribution of retained weights. An optional seventh panel displays correlations between the standardized PRS variables and ancestry principal components when the necessary aggregate table is present.

**Local PRS-decile and ND-CNV-stratified validation analyses**

A separate local analysis package was used to evaluate the relationship between the standardized schizophrenia PRS and observed neuropsychiatric phenotype burden across the analytic cohort. These analyses were performed from the ID-free SEM-ready dataset exported by Notebook 2 and therefore did not require access to participant identifiers or raw genomic data.

The source dataset contained 71,992 observations and included the standardized schizophrenia PRS variable `prs_scz_sem_z`, pooled recurrent ND-CNV carrier status (`nd_any`), broad psychosis status, and binary indicators for mood disorder, anxiety disorder, PTSD, autism, ADHD, intellectual disability, and developmental delay. An observed phenotype-burden score was calculated as the sum of these modeled psychiatric and developmental indicators. A binary elevated-burden variable was defined as the presence of two or more modeled phenotypes.

Participants were divided into ten approximately equal-sized groups according to the distribution of `prs_scz_sem_z`. Deciles contained 7,199 or 7,200 participants each. The package then generated four complementary summaries.

Option A estimated the prevalence of broad psychosis-spectrum phenotypes within each PRS decile. For each decile, the package reported the number of participants, number of psychosis cases, prevalence, and confidence intervals. Psychosis prevalence increased from 5.4% in the lowest decile to 6.7% in the highest decile.

Option B calculated the mean observed phenotype burden within each PRS decile. Outputs included the participant count, mean, standard deviation, standard error, and confidence interval. Mean burden increased from approximately 0.73 modeled phenotypes in the lowest decile to approximately 0.77 in the highest decile.

Option C calculated the prevalence of elevated multi-phenotype burden, defined as two or more modeled phenotypes, within each PRS decile. Elevated burden increased from 22.7% in the lowest decile to 24.2% in the highest decile.

Option D stratified psychosis prevalence jointly by PRS decile and recurrent ND-CNV carrier status. Within each decile, separate prevalence estimates and confidence intervals were calculated for recurrent ND-CNV carriers and non-carriers. The purpose of this visualization was to assess whether the carrier–non-carrier difference widened materially at higher levels of polygenic risk or remained relatively stable across the PRS distribution.

The accompanying provenance workbook additionally recorded a covariate-adjusted logistic model evaluating psychosis as a function of schizophrenia PRS, pooled recurrent ND-CNV carrier status, and their interaction. In this model, the schizophrenia PRS term was positively associated with psychosis risk, whereas the interaction term remained close to the null. The package also included a carrier-only exploratory regression of psychosis on PRS; this estimate was imprecise because the number of recurrent ND-CNV carriers within each decile was substantially smaller than the non-carrier sample.

The package includes:

- `prs_decile_analysis_table_no_ids.csv`, the ID-free analysis table containing PRS decile, standardized PRS, pooled ND-CNV status, individual phenotype indicators, total phenotype burden, and the elevated-burden indicator.
- `OptionA_psychosis_prevalence_by_prs_decile_source.csv`, containing psychosis prevalence and confidence intervals by PRS decile.
- `OptionB_mean_phenotype_burden_by_prs_decile_source.csv`, containing mean phenotype burden, standard deviation, standard error, and confidence intervals by PRS decile.
- `OptionC_multi_phenotype_burden_ge2_by_prs_decile_source.csv`, containing prevalence and confidence intervals for burden of two or more phenotypes.
- `OptionD_psychosis_prevalence_by_prs_decile_ndcnv_stratified_source.csv`, containing psychosis prevalence by PRS decile and pooled ND-CNV carrier status.
- Corresponding PNG and PDF figures for Options A–D.
- `PRS_decile_provenance_and_additional_analyses.xlsx`, containing a figure-provenance table, the joint PRS–ND-CNV interaction model, and the carrier-only exploratory PRS model.

Because the decile analyses were intended primarily as descriptive validation and visualization of the primary regression findings, interpretation focused on the overall gradient across the PRS distribution and the relative separation between carrier and non-carrier groups rather than on formal comparisons between individual deciles.

### IV. Code Description for the Lavaan CFA/SEM Pipeline

#### Overview of the CFA/SEM analysis workflow

This R Markdown pipeline performs confirmatory factor analysis and structural equation modeling to evaluate whether schizophrenia polygenic risk score, recurrent neurodevelopmental copy-number variant carrier status, and their interaction predict shared latent psychiatric and developmental liability in the *All of Us* Research Program cohort. The workflow uses the SEM-optimized participant-level export generated from the upstream *All of Us* PRS and ND-CNV logistic regression pipeline. The analysis is intentionally restricted to electronic health record-derived psychiatric and neurodevelopmental phenotypes. Survey-based dimensional modeling is not included because CDRv8 mental health survey availability, sample size, and partial-instrument structure currently limit interpretability.

The SEM hierarchy is organized around three conceptual models. Model 1 is the primary model and defines a broad latent psychiatric/developmental liability factor excluding psychosis. Model 2 treats psychosis as a downstream or specific phenotype predicted by broad latent psychiatric/developmental liability and by the genetic predictors. Model 3 is a sensitivity model that includes psychosis directly as an indicator of the latent factor. The primary genetic predictors are schizophrenia PRS, recurrent ND-CNV carrier status, and the PRS-by-ND-CNV interaction. Direction-specific exploratory models repeat the main framework using deletion-specific and duplication-specific ND-CNV carrier status.

#### Package loading and computational environment

The pipeline begins by defining and installing required R packages if they are missing. The core packages are `tidyverse`, `lavaan`, `semTools`, `broom`, `janitor`, `glue`, `readr`, `knitr`, and `kableExtra`. The `tidyverse` packages are used for data import, cleaning, reshaping, functional iteration, table construction, and plotting. The `lavaan` package is the main structural equation modeling engine. `semTools` provides additional SEM-related support functions, while `broom`, `janitor`, `glue`, `knitr`, and `kableExtra` support model output extraction, table formatting, and report generation.

The rendered HTML output reports that the SEM models were run using `lavaan` version 0.6-21. The model output uses a diagonally weighted least squares estimator, reported by lavaan as DWLS, corresponding to the WLSMV estimator requested in the code. This estimator is appropriate for ordered categorical indicators, which is important because the psychiatric and developmental outcomes are binary EHR-derived indicators rather than continuous measures.

#### User settings and expected input file

The user settings section defines the SEM input file and output directory. The expected input is `df_sem_lavaan_internal_no_ids.csv`, which is the ID-free SEM export produced by the upstream Notebook 2 workflow. This file is designed to contain one row per analytic participant and no row-level identifiers. The expected output directory is `sem_lavaan_outputs`, where model fit indices, factor loadings, structural paths, sensitivity outputs, and summary tables are written as CSV files.

The core input table must contain the primary schizophrenia PRS variable, recurrent ND-CNV exposure variables, interaction terms or variables sufficient to recreate those interaction terms, EHR-derived psychiatric indicators, and the covariates used in the SEMs. The primary PRS variable is `prs_scz_sem_z`. The main recurrent ND-CNV exposure is `nd_any`. Direction-specific exploratory exposures are `nd_del_any` and `nd_dup_any`. The main interaction term is `prs_x_nd_any`, and direction-specific interaction terms are `prs_x_nd_del` and `prs_x_nd_dup`.

**Main SEM predictors and covariates**

The primary predictor is the harmonized schizophrenia PRS variable `prs_scz_sem_z`. This variable represents the schizophrenia polygenic risk score after upstream ancestry-aware harmonization, residualization, and standardization in the AoUPRS workflow. The recurrent ND-CNV exposure is modeled as a binary indicator, `nd_any`, indicating whether a participant carries at least one recurrent neurodevelopmental CNV under the primary ro30 participant-level overlap definition. Direction-specific indicators, `nd_del_any` and `nd_dup_any`, indicate whether the participant carries at least one recurrent deletion or recurrent duplication, respectively.

The primary covariates are `age_last_obs`, `sex_at_birth_binary_centered`, `log1p_n_unique_visits`, `observation_time_days`, and genetic ancestry principal components `pc1` through `pc5`. Age at last observation captures participant age at the end of available EHR follow-up. The centered binary sex-at-birth covariate represents sex recoded into a numeric centered form suitable for regression and SEM. The log-transformed unique-visit count controls for healthcare utilization, which is important because participants with more healthcare encounters have more opportunities to receive diagnostic codes. Observation time controls for the amount of longitudinal EHR capture. Principal components PC1–PC5 adjust for genetic ancestry and population structure.

**Loading and validating the SEM dataframe**

The pipeline reads `df_sem_lavaan_internal_no_ids.csv` with `readr::read_csv` and prints the number of rows, number of columns, and a glimpse of the dataframe. In the rendered HTML output, the main SEM models use 71,992 observations and one missing-data pattern, indicating that the primary SEM dataset was complete for the modeled variables after preprocessing.

The code then defines the primary EHR-derived SEM indicators. The non-psychosis indicators are `mood_disorder`, `anxiety_disorder`, `ptsd`, `autism`, `adhd`, `intellectual_disability`, and `developmental_delay`. A second indicator set adds `psychosis_broad` to these indicators for models that either predict psychosis downstream or include psychosis directly in the measurement model.

The code uses a helper function, `recode_01`, to coerce binary indicators into clean integer 0/1 variables. This function explicitly recognizes common representations of false and true values, including "0", "0.0", "FALSE", "False", "false", "1", "1.0", "TRUE", "True", and "true". Values outside these expected encodings are set to missing. After recoding, the pipeline creates an indicator QC table summarizing the number of nonmissing participants, cases, controls, and unique values for each SEM indicator. The code stops if any indicator has no usable values, no cases, or no controls, preventing lavaan from fitting a model with noninformative binary outcomes.

**Predictor harmonization and compatibility with older SEM exports**

The harmonization section makes the R Markdown compatible with both the optimized current SEM export and older versions of the SEM CSV. The preferred PRS variable is `prs_scz_sem_z`, but the code includes fallback logic in case older files contain alternative PRS column names. This design prevents the notebook from failing simply because a prior export used a slightly different PRS variable name.

The code also harmonizes the sex covariate. The preferred sex covariate is `sex_at_birth_binary_centered`, but the notebook includes fallback behavior for older SEM exports where sex may have been stored under a less-optimized name or in a raw encoding. The goal is to ensure that sex enters lavaan as a numeric centered covariate rather than as an unprocessed OMOP concept ID or character variable.

The ND-CNV exposure flags are also checked and coerced into numeric binary variables. The code verifies that the main ND-CNV exposure variables exist and have usable values. It then recreates the PRS-by-CNV interaction terms from the current primary SEM PRS variable to avoid relying on stale interaction columns generated from older PRS variables. This step is important because if the PRS column changes, any previously exported interaction term may no longer correspond to the predictor actually used in the model. Recomputing interactions inside the SEM pipeline ensures internal consistency between `prs_scz_sem_z`, `nd_any`, and `prs_x_nd_any`.

**Definition and ordering of phenotype indicators**

The primary measurement model is built from seven non-psychosis EHR-derived indicators: mood disorder, anxiety disorder, PTSD, autism, ADHD, intellectual disability, and developmental delay. These indicators are used to define the general psychiatric/developmental latent factor in Model 1 and Model 2. The psychosis variable, `psychosis_broad`, is handled separately depending on the model. In Model 2, psychosis is modeled downstream of the latent factor. In Model 3, psychosis is included as an additional factor-loading indicator.

Before fitting lavaan models, the indicators are forced to ordered factors. This is required because the indicators are binary categorical outcomes and the code fits models using WLSMV estimation. Treating the outcomes as ordered categorical variables ensures that lavaan models the appropriate threshold-based measurement structure rather than assuming normally distributed continuous indicators.

**Descriptive quality-control outputs**

Before fitting SEM models, the notebook creates and exports descriptive QC summaries. The first table, `sem_indicator_counts.csv`, reports case counts, control counts, nonmissing counts, and prevalence for each SEM indicator. This output documents whether each binary phenotype has adequate variation for latent-variable modeling. The second table, `sem_exposure_counts.csv`, summarizes the number and prevalence of participants carrying any recurrent ND-CNV, recurrent deletion, and recurrent duplication. These outputs provide the sample-size and phenotype-distribution context needed to interpret the SEM models.

**Core SEM helper functions**

The pipeline defines several reusable helper functions to standardize model fitting and output extraction. `fit_sem_safely` takes lavaan model syntax, the SEM dataframe, a vector of ordered variables, and a model name, then attempts to fit the model with `lavaan::sem`. It uses the WLSMV estimator, theta parameterization, and pairwise missing-data handling. The fitting call is wrapped in `tryCatch`, so if a model fails, the notebook returns `NULL` and records a message rather than crashing the entire pipeline.

The `extract_fit` function extracts model-level fit indices, including convergence status, chi-square statistic, model degrees of freedom, p-value, comparative fit index, Tucker-Lewis index, root mean square error of approximation, and standardized root mean square residual. The `extract_paths` function extracts all lavaan parameter estimates with standardized estimates and confidence intervals. The `extract_structural_paths` function filters those estimates to regression-style paths with operator ~, which are the paths used to evaluate PRS, ND-CNV, interaction, covariate, and latent-factor effects. The `extract_loadings` function extracts factor loadings with operator `=~`, including standardized loadings and confidence intervals.

These helper functions make the output reproducible and consistent across the primary models, direction-specific exploratory models, leave-one-indicator-out models, CFA models, and bifactor-informed SEMs.

**Model syntax construction**

The notebook first defines functions that programmatically generate the lavaan syntax for the three main SEMs utilizing a generic one-factor SEM structure as proof of principle for each model and sensitivity output. Importantly, this section of the notebook is not used interpretively for the purpose of this publication. This design avoids duplicating long lavaan model strings and allows the same model structure to be reused with different ND-CNV exposures and interaction terms.

`make_model_1` defines a general psychiatric/developmental latent factor, `general_psych`, measured by mood disorder, anxiety disorder, PTSD, autism, ADHD, intellectual disability, and developmental delay. This latent factor is regressed on schizophrenia PRS, the selected ND-CNV exposure, the selected PRS-by-CNV interaction, age, centered sex, log-transformed visit count, observation time, and ancestry PCs 1–5. In the primary Model 1, the exposure is `nd_any` and the interaction is `prs_x_nd_any`.

`make_model_2` uses the same measurement and structural model for `general_psych` but adds psychosis as a downstream outcome. In this model, `psychosis_broad` is regressed on the latent factor, schizophrenia PRS, the selected ND-CNV exposure, the selected interaction term, and the same covariates. This model tests whether psychosis is better represented as a phenotype emerging downstream of broader psychiatric/developmental liability rather than being built directly into the latent factor.

`make_model_3` defines a broader measurement model in which `psychosis_broad` is included directly as an indicator of `general_psych`, together with mood disorder, anxiety disorder, PTSD, autism, ADHD, intellectual disability, and developmental delay. The resulting latent factor is then regressed on schizophrenia PRS, ND-CNV carrier status, the interaction term, and the same covariates. This model is used as a sensitivity analysis to test whether the conclusions change when psychosis is incorporated into the latent measurement structure.

**Model 1: Primary SEM excluding psychosis**

Model 1 is the primary SEM. It tests whether schizophrenia PRS, recurrent ND-CNV carrier status, and the PRS-by-ND-CNV interaction predict a broad psychiatric/developmental latent liability factor that excludes psychosis. The measurement portion of the model defines `general_psych` using mood disorder, anxiety disorder, PTSD, autism, ADHD, intellectual disability, and developmental delay. The structural portion regresses `general_psych` on `prs_scz_sem_z`, `nd_any`, `prs_x_nd_any`, age, sex, utilization, observation time, and ancestry PCs.

The code fits the model with `lavaan::sem`, using WLSMV estimation, theta parameterization, ordered binary indicators, and pairwise missing-data handling. The model summary prints fit measures, standardized estimates, and R-squared values. After fitting, the notebook extracts three outputs: model fit indices, factor loadings, and structural paths. These are written to `model_1_fit_indices.csv`, `model_1_factor_loadings.csv`, and `model_1_structural_paths.csv`.

The logic of Model 1 is to estimate whether schizophrenia-associated common and rare genetic liability contributes to broad psychiatric/developmental liability independent of psychosis placement. Because psychosis is excluded from the measurement model, this model avoids defining the latent factor around the outcome most directly associated with schizophrenia PRS.

**Model 2: Psychosis as a downstream phenotype**

Model 2 retains the same non-psychosis latent factor but explicitly models psychosis as a downstream phenotype. The `general_psych` factor is again measured by mood disorder, anxiety disorder, PTSD, autism, ADHD, intellectual disability, and developmental delay. The latent factor is regressed on schizophrenia PRS, ND-CNV carrier status, the PRS-by-ND-CNV interaction, and covariates. Separately, `psychosis_broad` is regressed on the latent factor, PRS, ND-CNV carrier status, the interaction term, and the same covariates.

This model requires `psychosis_broad` to be present in the SEM dataset. If the psychosis column is absent, the code skips Model 2 and records a message. In the attached run, `psychosis_broad` was present and Model 2 was fitted successfully.

Model 2 is conceptually important because it tests whether broad psychiatric/developmental liability explains psychosis-spectrum phenotypes while also allowing PRS and ND-CNV carrier status to have direct effects on psychosis. This avoids circularity because psychosis is not used to define the latent factor that then predicts psychosis. The model produces `model_2_fit_indices.csv`, `model_2_factor_loadings.csv`, and `model_2_structural_paths.csv`.

**Model 3: Psychosis included in the latent factor**

Model 3 is a sensitivity model in which psychosis is included directly as an indicator of the general psychiatric/developmental latent factor. The measurement model includes mood disorder, anxiety disorder, PTSD, psychosis broad, autism, ADHD, intellectual disability, and developmental delay. The latent factor is then regressed on schizophrenia PRS, ND-CNV carrier status, the PRS-by-ND-CNV interaction, and covariates.

The purpose of Model 3 is to test whether the overall conclusions are sensitive to treating psychosis as part of the latent measurement structure rather than as a downstream phenotype. Like the other models, it uses WLSMV estimation, theta parameterization, ordered binary indicators, and pairwise missing-data handling. The code wraps model fitting in `tryCatch` so that model failure returns a controlled message rather than interrupting the notebook. Model 3 outputs are saved as `model_3_fit_indices.csv`, `model_3_factor_loadings.csv`, and `model_3_structural_paths.csv`.

**Direction-specific exploratory SEMs**

After fitting the any-ND-CNV primary models, the pipeline runs direction-specific exploratory SEMs using deletion and duplication carrier status. These models use the Model 1 framework, excluding psychosis from the latent factor, but replace `nd_any` and `prs_x_nd_any` with either `nd_del_any` and `prs_x_nd_del` or `nd_dup_any` and `prs_x_nd_dup`.

The code defines a `directional_specs` table listing the model names, exposure variables, and interaction variables for deletion- and duplication-specific models. It then uses `pmap` to iterate over the model specifications, fit each model safely, and extract fit indices, loadings, and structural paths. The outputs are combined into three tables: `direction_specific_fit_indices.csv`, `direction_specific_factor_loadings.csv`, and `direction_specific_structural_paths.csv`.

These models are exploratory because deletion and duplication carrier groups are smaller than the pooled ND-CNV group. The notebook explicitly notes that interaction terms should be interpreted cautiously because direction-specific analyses are less powered than the primary `nd_any` models.

**Leave-one-indicator-out sensitivity analyses**

The leave-one-indicator-out analysis evaluates whether the primary latent factor is driven by any single EHR-derived phenotype. The code defines `make_leave_one_out_model`, which rebuilds the Model 1 lavaan syntax after removing one indicator from the measurement model. For each indicator in the non-psychosis indicator set, the model is refit using the remaining indicators, and the same predictors and covariates are retained.

The code uses safe helper functions to extract parameter estimates and fit indices even when individual models fail or return unexpected lavaan objects. For each leave-one-out model, the pipeline records whether the model converged, its fit indices, structural paths, and factor loadings. The primary interpretation is whether the PRS, ND-CNV, and interaction paths remain directionally stable after removing each indicator. If the genetic path estimates remain similar across leave-one-out models, this supports the conclusion that the latent structure is not dominated by a single high-prevalence phenotype.

**CFA comparison of empirical latent structure**

The CFA section evaluates measurement structure before interpreting genetic paths. It compares whether the EHR indicators are better represented as one broad psychiatric/developmental factor, two correlated factors, or a bifactor structure. This is a measurement-only step: no PRS, CNV exposure, interaction term, or covariates are included.

The one-factor CFA model defines a single latent factor loading on all EHR indicators. The correlated two-factor model separates internalizing and neurodevelopmental indicators into two correlated latent factors. The bifactor model defines a general factor loading on all indicators and domain-specific factors loading on subsets of indicators. The bifactor model also constrains the general and domain-specific factors to be orthogonal, meaning their covariances are fixed to zero. This allows the general factor to represent shared liability across indicators while the specific factors absorb residual covariance within internalizing or neurodevelopmental domains.

The CFA models are fit safely using WLSMV estimation for ordered binary indicators. The code extracts fit indices, standardized factor loadings, and factor correlations where available. The exported CFA outputs are `lavaan_cfa_fit_indices.csv`, `lavaan_cfa_loadings.csv`, and `lavaan_cfa_factor_correlations.csv`. These tables support the selection of bifactor-informed SEM sensitivity models.

**Bifactor-informed SEM sensitivity models**

The bifactor-informed SEM section extends the CFA comparison into genetic path modeling. These models preserve the substantive logic of the original M1–M3 hierarchy while adding domain-specific residual factors suggested by the CFA results. The general factor remains the primary target of interpretation. The specific factors are included to account for residual covariance within symptom domains rather than to serve as primary genetic outcomes.

The bifactor Model 1 defines a general psychiatric/developmental factor excluding psychosis and includes internalizing-specific and neurodevelopmental-specific factors. The general factor loads on mood disorder, anxiety disorder, PTSD, autism, ADHD, intellectual disability, and developmental delay. The internalizing-specific factor loads on mood disorder, anxiety disorder, and PTSD. The neurodevelopmental-specific factor loads on autism, ADHD, intellectual disability, and developmental delay. The general factor, internalizing-specific factor, and neurodevelopmental-specific factor are constrained to be orthogonal. The general factor is regressed on schizophrenia PRS, ND-CNV carrier status, PRS-by-ND-CNV interaction, and covariates. This model is saved as the bifactor Model 1 output.

The bifactor Model 2 uses the same bifactor measurement structure but models psychosis as a downstream phenotype. In this model, `psychosis_broad` is regressed on the general latent factor, PRS, ND-CNV carrier status, the interaction term, and covariates. This model tests whether psychosis is predicted by broad bifactor-derived psychiatric/developmental liability while preserving the distinction between general and domain-specific covariance.

The bifactor Model 3 includes psychosis directly in the bifactor measurement structure. This model tests whether conclusions change when psychosis contributes directly to the general factor while retaining internalizing- and neurodevelopmental-specific residual factors. Because bifactor models with sparse binary indicators can be fragile, all bifactor models are fit with guarded error handling and interpreted with attention to convergence and residual fit.

The bifactor Model 1 outputs are `bifactor_model_1_fit_indices.csv`, `bifactor_model_1_factor_loadings.csv`, and `bifactor_model_1_structural_paths.csv`. The bifactor Model 2 outputs are `bifactor_model_2_fit_indices.csv`, `bifactor_model_2_factor_loadings.csv`, and `bifactor_model_2_structural_paths.csv`. The bifactor Model 3 outputs are `bifactor_model_3_fit_indices.csv`, `bifactor_model_3_factor_loadings.csv`, and `bifactor_model_3_structural_paths.csv`.

**Bifactor direction-specific exploratory SEMs**

The pipeline also implements bifactor-informed deletion- and duplication-specific exploratory SEMs. These models replace the pooled ND-CNV exposure with either deletion carrier status or duplication carrier status while retaining the bifactor measurement architecture. The purpose is to assess whether the general psychiatric/developmental liability paths differ when rare structural variation is separated by CNV direction.

The direction-specific bifactor outputs include `bifactor_direction_specific_fit_indices.csv`, `bifactor_direction_specific_factor_loadings.csv`, `bifactor_direction_specific_structural_paths.csv`, and `bifactor_direction_specific_key_paths.csv`. The key-path output is particularly useful for manuscript figures because it extracts the most interpretable paths, such as schizophrenia PRS to general liability, ND-CNV exposure to general liability, PRS-by-CNV interaction to general liability, and latent liability to psychosis where applicable.

**Bifactor leave-one-indicator-out sensitivity analysis**

The bifactor leave-one-indicator-out section repeats the leave-one-indicator-out logic within the bifactor measurement framework. For each indicator, the code removes that indicator from the general factor and from any relevant domain-specific factor, rebuilds the bifactor model syntax, refits the model, and extracts fit indices, structural paths, factor loadings, and key paths.

This analysis evaluates whether the bifactor-derived general liability factor and genetic path estimates are stable when individual phenotypes are removed. Because some indicators are sparse, especially autism, intellectual disability, and developmental delay, this sensitivity analysis is important for showing whether the model is robust to rare-outcome behavior and not driven entirely by common affective phenotypes. The outputs are `bifactor_leave_one_indicator_out_fit_indices.csv`, `bifactor_leave_one_indicator_out_structural_paths.csv`, `bifactor_leave_one_indicator_out_factor_loadings.csv`, and `bifactor_leave_one_indicator_out_key_paths.csv`.

**Combined bifactor SEM summary tables**

After fitting the bifactor models and sensitivities, the pipeline creates combined summary tables. These tables aggregate fit indices, structural paths, and key interpretable paths across bifactor Model 1, bifactor Model 2, bifactor Model 3, direction-specific bifactor models, and bifactor leave-one-out models.

The main combined outputs are `bifactor_sem_fit_summary.csv`, `bifactor_sem_structural_paths.csv`, and `bifactor_sem_key_paths.csv`. The key-path table is designed to support interpretation and visualization by reducing the full lavaan parameter table to the subset of paths most relevant to the scientific question. These include PRS effects, ND-CNV effects, interaction effects, and latent liability effects on psychosis.

**Primary interpretation helper table**

The primary interpretation helper section extracts and organizes the model paths most relevant for manuscript interpretation. The goal is to avoid forcing readers to interpret full lavaan parameter tables containing every loading, threshold, variance, covariance, and covariate path. Instead, the helper table focuses on the genetic and latent-structure paths that correspond to the manuscript hypotheses.

For Model 1, the key paths are PRS to general psychiatric/developmental liability, ND-CNV carrier status to general liability, and PRS-by-ND-CNV interaction to general liability. For Model 2, the key paths include those same paths plus general liability to psychosis and direct PRS, ND-CNV, and interaction effects on psychosis. For Model 3, the key paths again focus on PRS, ND-CNV, and interaction effects on the general latent factor, now with psychosis included as a measurement indicator.

This section is useful for manuscript writing because it directly supports statements about additive versus interactive genetic architecture, whether psychosis is better modeled downstream of broader liability, and whether genetic effects remain stable across alternative psychosis placements.

#### Diagnostic and publication plotting

The final major analytic section generates a comprehensive plot suite from the exported SEM and CFA tables. The plot directory is `sem_plots`. The code creates `primary_sem_structural_paths.png`, which visualizes standardized structural paths for primary SEM models. It also creates `sem_model_fit_comparison.png`, which compares fit indices across SEM models, and `cfa_fit_comparison.png`, which compares CFA measurement structures.

The code additionally generates `cfa_standardized_loadings.png`, which visualizes standardized factor loadings across CFA models, and `leave_one_indicator_out_key_paths.png`, which summarizes the stability of key structural paths across leave-one-indicator-out sensitivity models. If direction-specific SEM path tables are available, the notebook also generates `direction_specific_sem_paths.png`, showing deletion- and duplication-specific structural paths.

These plots support both internal model diagnostics and manuscript figure development. They allow the analyst to compare model fit, inspect whether indicators load in expected directions, evaluate whether key genetic paths remain stable across sensitivity models, and visualize deletion-versus-duplication differences.

#### Complete input and output inventory

The primary input is `df_sem_lavaan_internal_no_ids.csv`, an ID-free participant-level SEM dataset exported from the upstream *All of Us* PRS and ND-CNV modeling notebook. This file must contain EHR-derived binary phenotype indicators, schizophrenia PRS, ND-CNV exposure variables, interaction terms or source variables for recreating them, and covariates. The required phenotype indicators are `mood_disorder`, `anxiety_disorder`, `ptsd`, `autism`, `adhd`, `intellectual_disability`, `developmental_delay`, and, for psychosis models, `psychosis_broad`. The required genetic variables are `prs_scz_sem_z`, `nd_any`, `nd_del_any`, and `nd_dup_any`. The required covariates are `age_last_obs`, `sex_at_birth_binary_centered`, `log1p_n_unique_visits`, `observation_time_days`, and ancestry PCs `pc1` through `pc5`.

The main output directory, `sem_lavaan_outputs`, contains QC tables, model fit tables, factor loading tables, structural path tables, CFA outputs, bifactor SEM outputs, direction-specific outputs, leave-one-out outputs, and combined interpretation tables. The plot output directory, `sem_plots`, contains model diagnostic and manuscript-supporting figures.

The QC outputs are `sem_indicator_counts.csv` and `sem_exposure_counts.csv`. The primary SEM outputs are `model_1_fit_indices.csv`, `model_1_factor_loadings.csv`, `model_1_structural_paths.csv`, `model_2_fit_indices.csv`, `model_2_factor_loadings.csv`, `model_2_structural_paths.csv`, `model_3_fit_indices.csv`, `model_3_factor_loadings.csv`, and `model_3_structural_paths.csv`. The direction-specific SEM outputs are `direction_specific_fit_indices.csv`, `direction_specific_factor_loadings.csv`, and `direction_specific_structural_paths.csv`. The CFA outputs are `lavaan_cfa_fit_indices.csv`, `lavaan_cfa_loadings.csv`, and lavaan_cfa_factor_correlations.csv. The bifactor outputs are bifactor_model_1_fit_indices.csv, bifactor_model_1_factor_loadings.csv, bifactor_model_1_structural_paths.csv, bifactor_model_2_fit_indices.csv, bifactor_model_2_factor_loadings.csv, bifactor_model_2_structural_paths.csv, bifactor_model_3_fit_indices.csv, bifactor_model_3_factor_loadings.csv, and bifactor_model_3_structural_paths.csv. The bifactor direction-specific outputs are bifactor_direction_specific_fit_indices.csv, bifactor_direction_specific_factor_loadings.csv, bifactor_direction_specific_structural_paths.csv, and bifactor_direction_specific_key_paths.csv. The bifactor leave-one-out outputs are bifactor_leave_one_indicator_out_fit_indices.csv, bifactor_leave_one_indicator_out_structural_paths.csv, bifactor_leave_one_indicator_out_factor_loadings.csv, and bifactor_leave_one_indicator_out_key_paths.csv. The combined bifactor summary outputs are bifactor_sem_fit_summary.csv, bifactor_sem_structural_paths.csv, and bifactor_sem_key_paths.csv.

**Reproducibility logic of the SEM pipeline**

A researcher seeking to reproduce this SEM pipeline would first need the SEM-ready export generated by the upstream *All of Us* PRS and ND-CNV notebook. That export must contain binary EHR-derived psychiatric and developmental indicators, schizophrenia PRS, recurrent ND-CNV carrier status variables, PRS-by-CNV interaction terms or source variables, demographic covariates, utilization covariates, observation-time covariates, and ancestry principal components. The researcher would then run the R Markdown file in an R environment with lavaan, semTools, and tidyverse-compatible packages installed.

The pipeline first validates the input dataset, cleans binary indicators, harmonizes predictors and covariates, recreates interaction terms, and converts indicators into ordered factors. It then fits primary SEM models testing three alternative placements of psychosis: excluded from the latent factor, modeled downstream of the latent factor, and included directly as a latent-factor indicator. It repeats the primary framework using deletion- and duplication-specific exposures, performs leave-one-indicator-out sensitivity analyses, compares empirical measurement structures using CFA, fits bifactor-informed SEM sensitivity models, summarizes key paths, and generates diagnostic plots.

The central modeling logic is that broad psychiatric and developmental liability can be represented as a latent variable inferred from multiple EHR-derived indicators. Schizophrenia PRS, recurrent ND-CNV carrier status, and their interaction are modeled as predictors of that latent liability. Psychosis is then evaluated under alternative assumptions: as excluded from the latent definition, as a downstream phenotype predicted by broad liability, or as an indicator of the latent factor itself. By comparing these models, the pipeline evaluates whether schizophrenia-associated genetic risk is better interpreted as acting through broader psychiatric/developmental liability dimensions rather than exclusively through psychosis-specific structure.

No false-discovery-rate q-values are calculated within this SEM R Markdown file. The SEM outputs instead report model fit indices, standardized factor loadings, regression path estimates, standard errors, z statistics, p values, confidence intervals, and standardized coefficients. This is appropriate because the SEM pipeline is organized around a small number of pre-specified latent-variable models and sensitivity analyses rather than a large phenotype-wide multiple-testing scan.

### V. Supplemental Figures

A

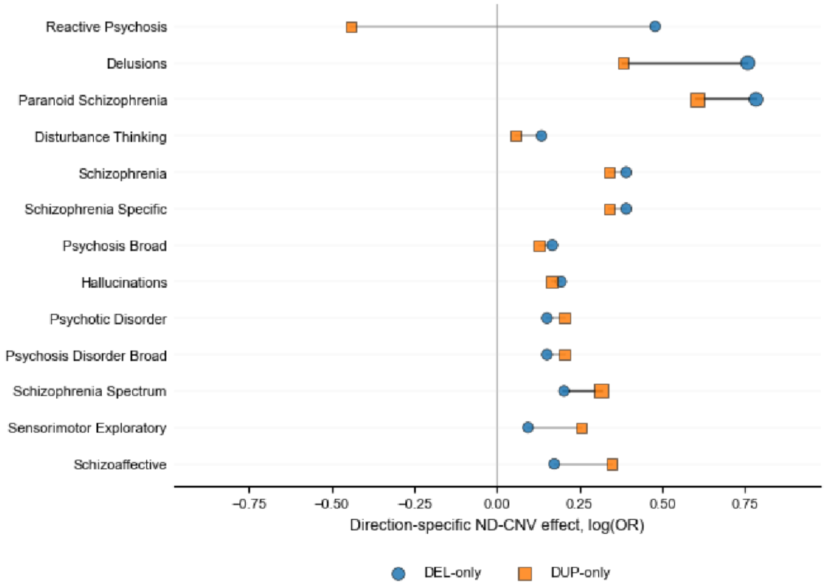


B

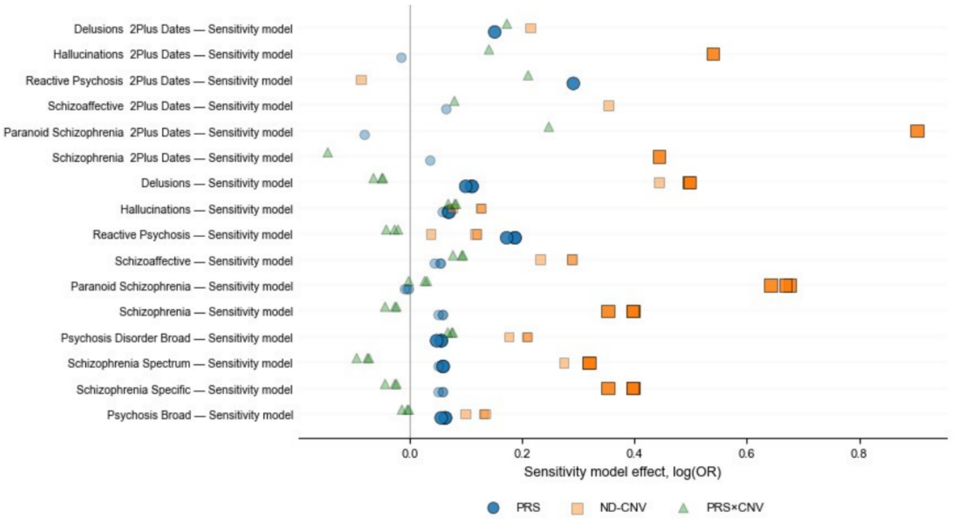


**Figure S1. Psychosis-spectrum association profiles and phenotype-definition sensitivity analyses. (A)** Phenotype-level direction-specific recurrent ND-CNV association profiles across deletion-specific (DEL), duplication-specific (DUP), and pooled ND-CNV carrier status models. Dot size reflects relative effect magnitude, while color intensity reflects standardized association direction and strength across modeled psychosis-spectrum and neurodevelopmental phenotypes. Panels display pooled, deletion-overlapping, and duplication-overlapping phenotype-level association profiles. **(B)** Effect estimates from primary regression models and alternate case-definition sensitivity models are shown for schizophrenia polygenic risk score (PRS), recurrent ND-CNV carrier status, and PRS × ND-CNV interaction terms across psychosis-spectrum phenotypes. Points represent log odds ratio estimates from the primary and alternate phenotype definitions requiring two or more qualifying diagnostic encounters ("2Plus Dates").

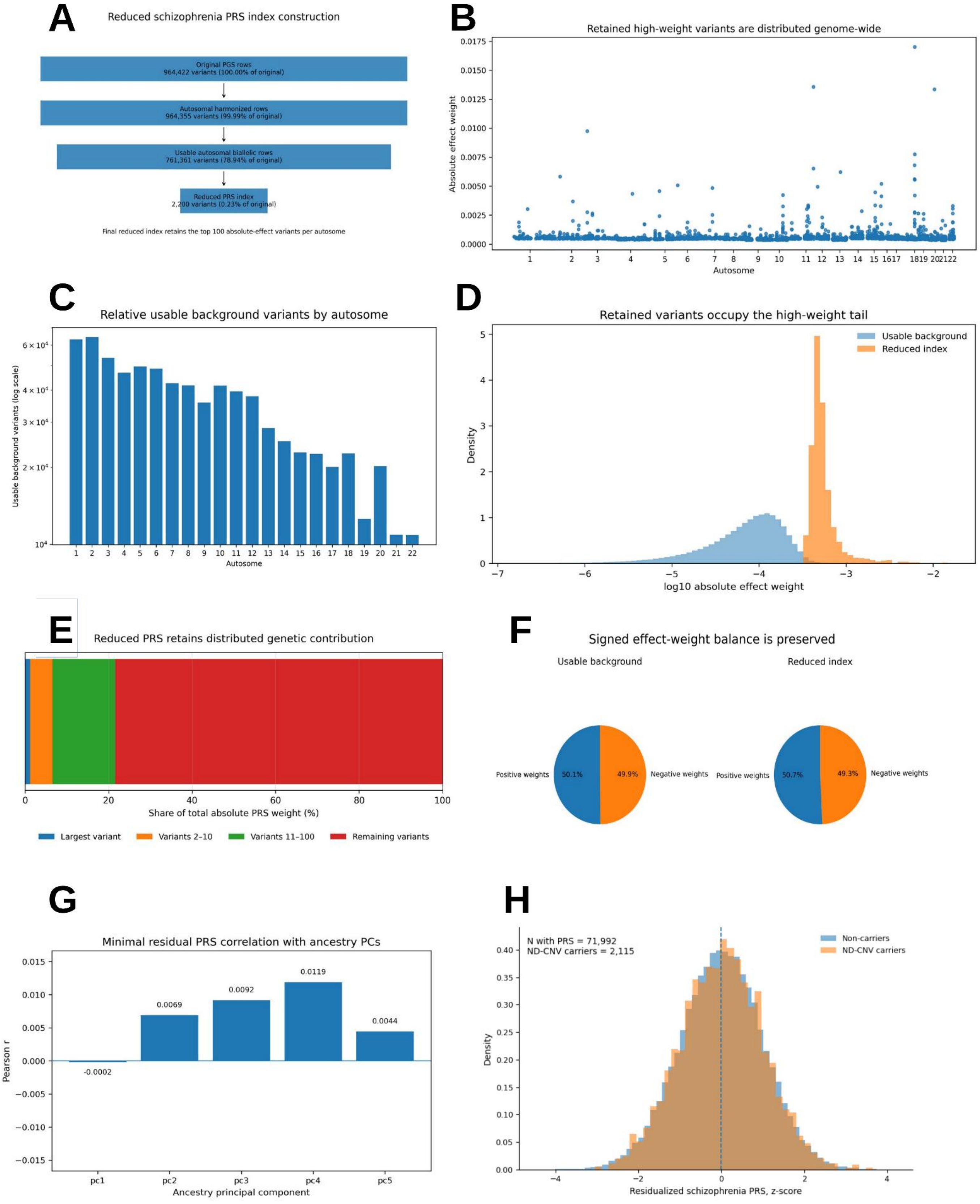
A
Reduced schizophrenia PRS index construction
Original PGS rows
964,422 variants (100.00% of original)
Autosomal harmonized rows
964,355 variants (99.99% of original)
Usable autosomal biallelic rows
761,361 variants (78.94% of original)
Reduced PRS index
2,200 variants (0.23% of original)
Final reduced index retains the top 100 absolute-effect variants per autosome
B
Retained high-weight variants are distributed genome-wide
Absolute effect weight
Autosome
C
Relative usable background variants by autosome
Usable background variants (log scale)
Autosome
D
Retained variants occupy the high-weight tail
Usable background
Reduced index
Density
log10 absolute effect weight
E
Reduced PRS retains distributed genetic contribution
Share of total absolute PRS weight (%)
Largest variant
Variants 2–10
Variants 11–100
Remaining variants
F
Signed effect-weight balance is preserved
Usable background
Reduced index
Positive weights
50.1%
49.9%
Negative weights
Positive weights
50.7%
49.3%
Negative weights
G
Minimal residual PRS correlation with ancestry PCs
Pearson r
-0.0002
0.0069
0.0092
0.0119
0.0044
pc1
pc2
pc3
pc4
pc5
Ancestry principal component
H
N with PRS = 71,992
ND-CNV carriers = 2,115
Non-carriers
ND-CNV carriers
Density
Residualized schizophrenia PRS, z-score

**Figure S2. Validation of the reduced schizophrenia polygenic risk score (PRS). (A)** Schematic overview of reduced schizophrenia PRS construction. The original PGS002785 schizophrenia PRS underwent quality-control filtering to generate a set of usable variants, after which the 100 variants with the largest absolute effect weights were retained from each autosome. This procedure yielded a final reduced score containing 2,200 variants (100 per autosome) while preserving genome-wide representation. **(B)** Genomic position and effect-weight magnitude of variants retained in the reduced PRS. Retained variants remained broadly distributed across all autosomes. **(C)** Relative number of usable background variants by autosome prior to score reduction. **(D)** Distribution of absolute variant effect weights in the usable background variant set and the reduced PRS. The reduced PRS was enriched for variants with larger absolute effect weights. **(E)** Distribution of absolute PRS weight contribution across variants retained in the reduced PRS. Variants are grouped according to their relative contribution to total absolute score weight, including the largest-weight variant, variants ranked 2–10, variants ranked 11–100, and all remaining retained variants. The largest retained variant contributed only 1.21% of total absolute PRS weight, while the top 10 and top 100 retained variants collectively contributed 6.55% and 21.58%, respectively. Despite selection based on absolute effect size, contribution to the reduced score remained distributed across many loci rather than being dominated by a small number of highly weighted variants. The reduced score demonstrated an effective variant count ($N_{eff}$) of 893.7, where $N_{eff}$ represents the number of equally weighted variants that would produce the same degree of weight concentration as the observed score. **(F)** Preservation of signed effect-weight balance following score reduction. Positive-effect and negative-effect variants remained nearly identical between the usable background variant set (50.15% positive, 49.85% negative) and the final reduced PRS (50.73% positive, 49.27% negative). **(G)** Correlation of reduced PRS values with the first ten genetic ancestry principal components. Correlation coefficients were uniformly small across all examined principal components. **(H)** Residualized schizophrenia polygenic risk score (PRS) distributions across recurrent ND-CNV carriers and non-carriers following ancestry-aware normalization and residualization procedures implemented through the AoUPRS framework. Residualized PRS distributions remained broadly centered near zero across the analytic cohort.

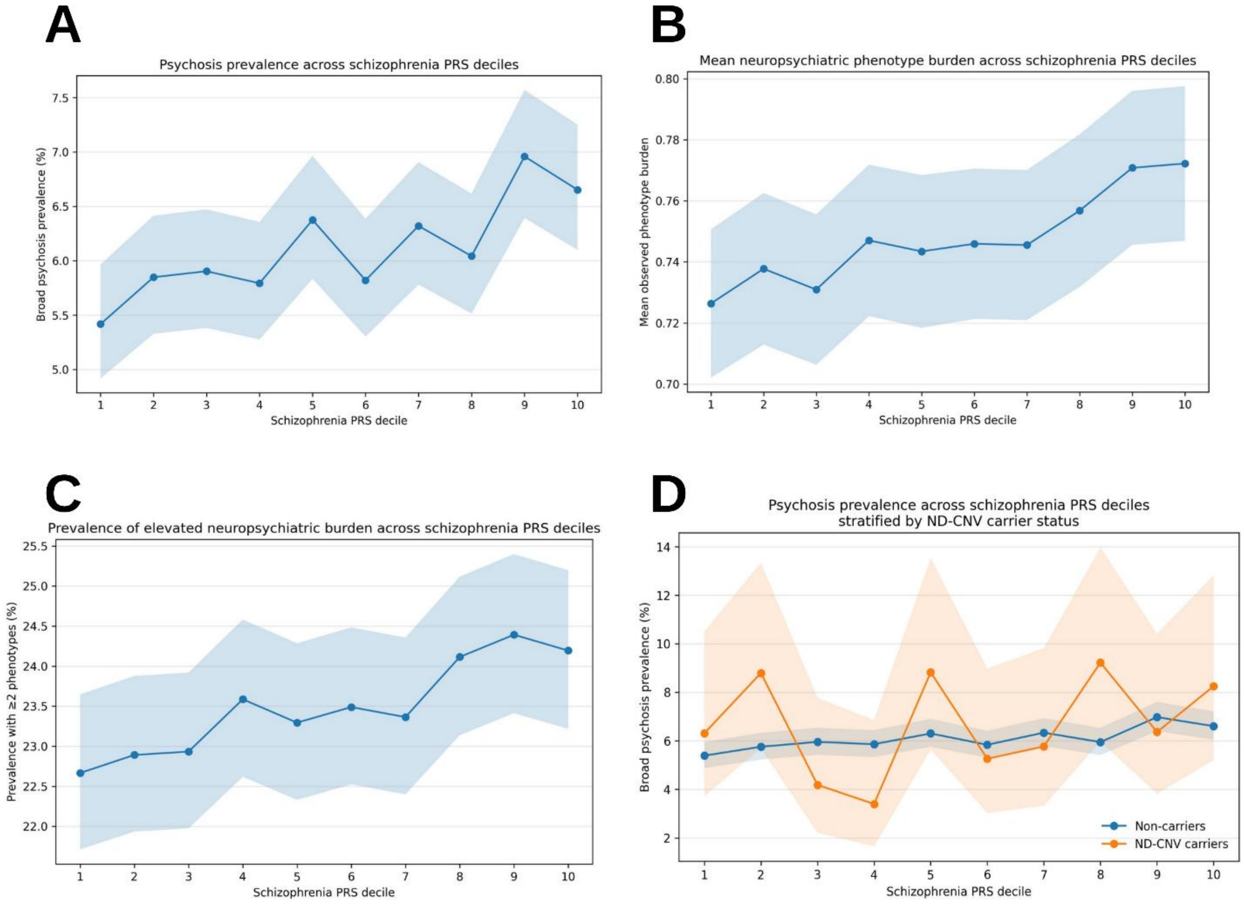


**Figure S3. Schizophrenia polygenic risk score deciles show graded psychosis-spectrum prevalence and neuropsychiatric phenotype burden. (A)** Psychosis-spectrum prevalence across schizophrenia PRS deciles. Prevalence increased from 5.4% in the lowest decile to 6.7% in the highest decile, corresponding to a 22.8% relative increase across the PRS distribution. **(B)** Mean observed neuropsychiatric phenotype burden across schizophrenia PRS deciles. Mean burden increased from 0.73 to 0.77 phenotypes across deciles (6.3% relative increase). **(C)** Prevalence of elevated neuropsychiatric burden, defined as two or more modeled phenotypes, across schizophrenia PRS deciles. Elevated burden prevalence increased from 22.7% to 24.2% (6.7% relative increase). **(D)** Psychosis-spectrum prevalence across schizophrenia PRS deciles stratified by recurrent ND-CNV carrier status. Psychosis prevalence increased across PRS strata in both carrier and non-carrier groups, while the separation between groups remained relatively stable across the PRS distribution.

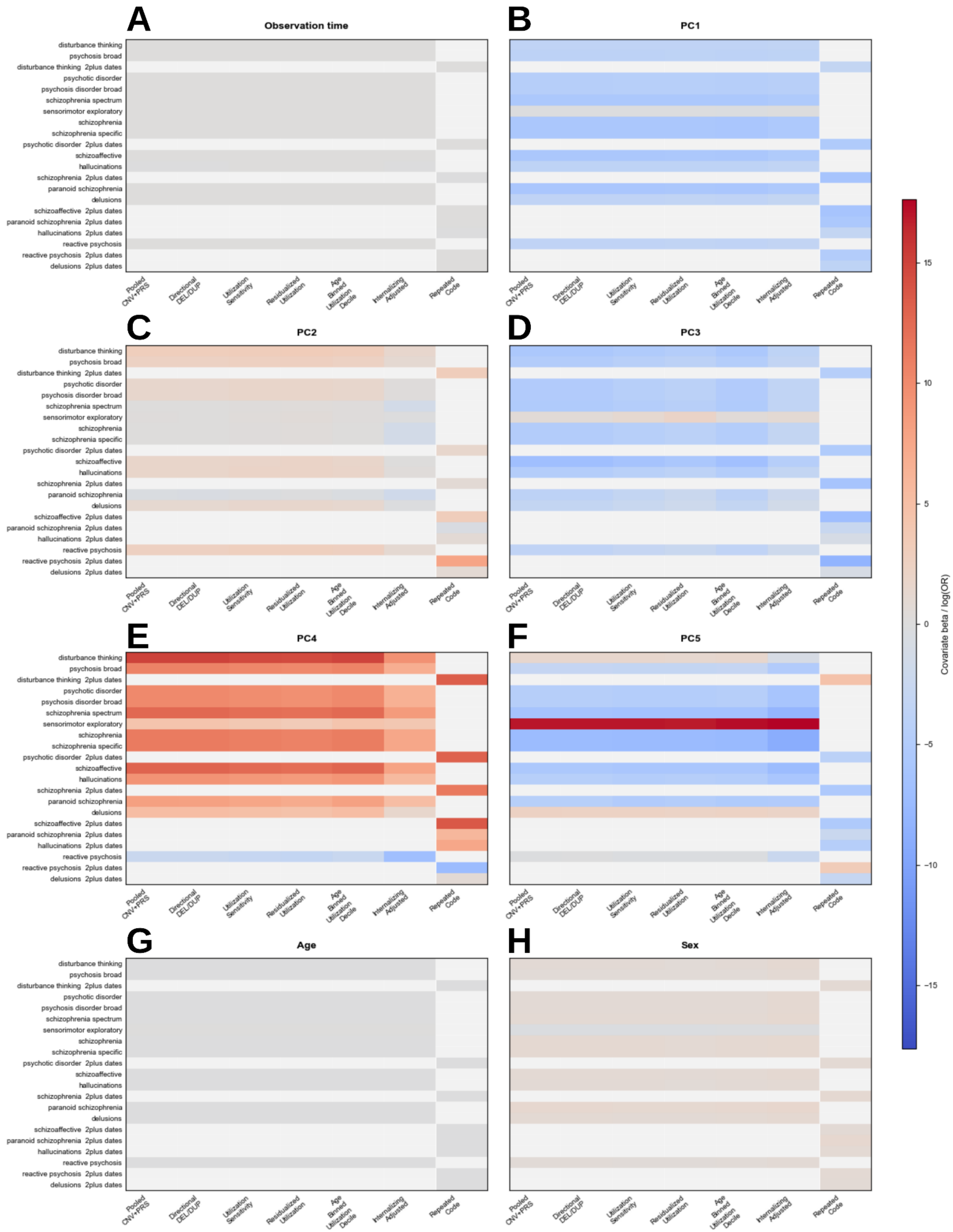

A
Observation time
B
PC1
C
PC2
D
PC3
E
PC4
F
PC5
G
Age
H
Sex
disturbance thinking
psychosis broad
disturbance thinking 2plus dates
psychotic disorder
psychosis disorder broad
schizophrenia spectrum
sensorimotor exploratory
schizophrenia
schizophrenia specific
psychotic disorder 2plus dates
schizoaffective
hallucinations
schizophrenia 2plus dates
paranoid schizophrenia
delusions
schizoaffective 2plus dates
paranoid schizophrenia 2plus dates
hallucinations 2plus dates
reactive psychosis
reactive psychosis 2plus dates
delusions 2plus dates
Pooled CNV+PRS
Directional DEL/DUP
Utilization Sensitivity
Residualized Utilization
Age Binned Utilization Decile
Internalizing Adjusted
Repeated Code
Covariate beta / log(OR)
15
10
5
0
−5
−10
−15

**Figure S4. Covariate perturbation analyses of phenotype-level regression models. (A-H)** Heatmaps summarizing covariate perturbation effects across schizophrenia PRS, recurrent ND-CNV carrier status, and PRS × ND-CNV interaction regression models. Panels correspond to alternate demographic, ancestry, healthcare-utilization, and longitudinal ascertainment covariate structures, including age, sex, ancestry principal components (PC1-PC5), healthcare-utilization burden, and observation duration perturbations. Color intensity reflects relative perturbation magnitude of standardized phenotype-level effect estimates across psychosis-spectrum and broader psychiatric phenotypes.

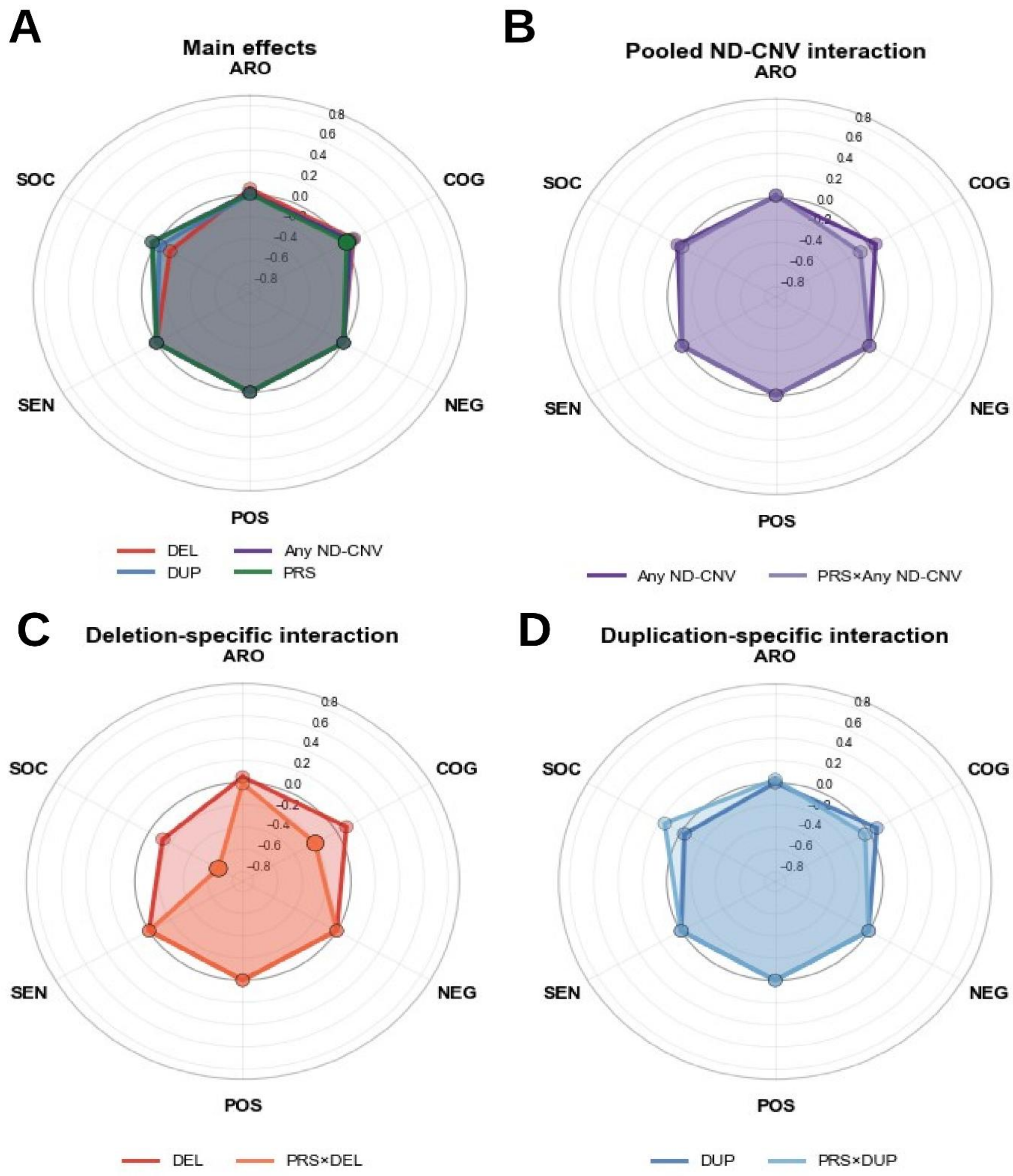


**Figure S5. Research Domain Criteria (RDoC)-informed dimensional summaries of the primary phenotype-level genetic association analyses. (A–D)** Radar plots summarize standardized associations across six RDoC-informed domains—cognitive systems, negative valence systems, positive valence systems, social processes, arousal/regulatory systems, and sensorimotor systems—for pooled recurrent ND-CNV carrier status (A), schizophrenia polygenic risk score (PRS) (B), deletion-overlapping recurrent ND-CNV carrier status (C), and duplication-overlapping recurrent ND-CNV carrier status (D). These complementary dimensional summaries recapitulate the primary phenotype-level association patterns observed in Figure 2.

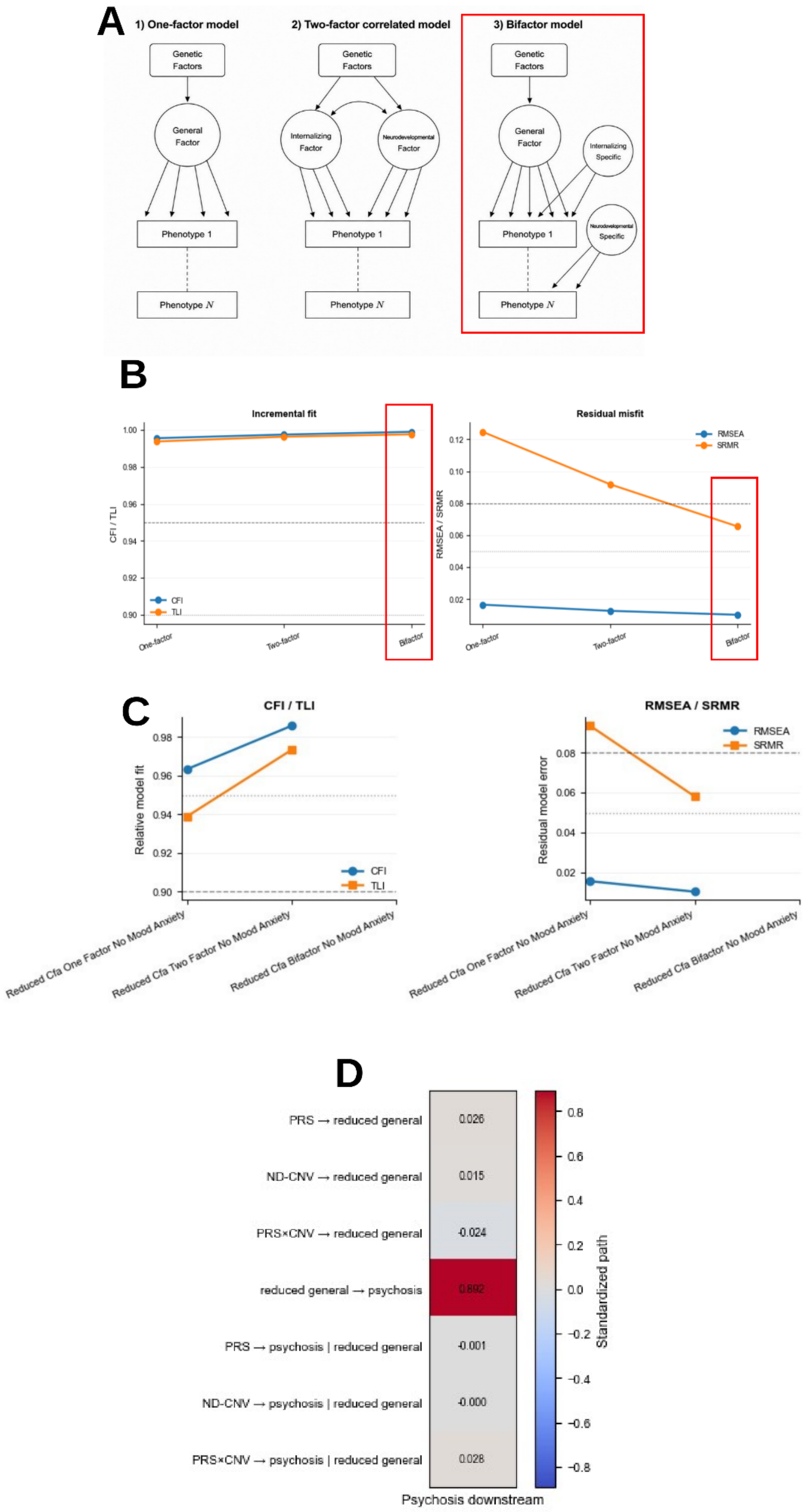
A
1) One-factor model
2) Two-factor correlated model
3) Bifactor model
Genetic Factors
General Factor
Internalizing Factor
Neurodevelopmental Factor
Internalizing Specific
Neurodevelopmental Specific
Phenotype 1
Phenotype N
B
Incremental fit
CFI / TLI
CFI
TLI
One-factor
Two-factor
Bifactor
Residual misfit
RMSEA / SRMR
RMSEA
SRMR
C
CFI / TLI
Relative model fit
RMSEA / SRMR
Residual model error
Reduced Cfa One Factor No Mood Anxiety
Reduced Cfa Two Factor No Mood Anxiety
Reduced Cfa Bifactor No Mood Anxiety
D
PRS → reduced general
ND-CNV → reduced general
PRS×CNV → reduced general
reduced general → psychosis
PRS → psychosis | reduced general
ND-CNV → psychosis | reduced general
PRS×CNV → psychosis | reduced general
0.026
0.015
-0.024
0.892
-0.001
-0.000
0.028
Standardized path
Psychosis downstream

**Figure S6. Confirmatory factor analysis and structural equation modeling sensitivity analyses.** **(A)** Minimalist schematic representations of confirmatory factor analysis (CFA) covariance models evaluated across psychiatric and neurodevelopmental phenotypes. Models included one-factor, correlated two-factor, and bifactor frameworks. Red highlighting denotes selection of the bifactor framework for downstream structural equation modeling analyses. **(B)** Comparative model fit and residual misfit estimates across one-factor, correlated two-factor, and bifactor CFA structures. Panels summarize comparative fit index (CFI), Tucker-Lewis index (TLI), root mean square error of approximation (RMSEA), and standardized root mean square residual (SRMR). Bifactor models showed the highest relative fit (CFI = 0.999, TLI = 0.998) and lowest residual misfit (RMSEA = 0.010, SRMR = 0.065), compared with correlated two-factor (RMSEA = 0.013, SRMR = 0.092) and one-factor alternatives (RMSEA = 0.017, SRMR = 0.125). **(C)** Comparative CFA fit estimates following exclusion of mood and anxiety phenotypes from the latent structure. Panels summarize CFI, TLI, RMSEA, and SRMR across reduced-factor one-factor, correlated two-factor, and bifactor frameworks. Following removal of highly prevalent affective phenotypes, the reduced-factor bifactor model failed to converge, whereas the correlated two-factor model demonstrated the strongest converged fit (CFI = 0.986, TLI = 0.973, RMSEA = 0.010, SRMR = 0.058). **(D)** Reduced-factor structural equation modeling analyses following exclusion of mood and anxiety phenotypes. Heatmaps summarize standardized path coefficients following exclusion of mood and anxiety phenotypes.

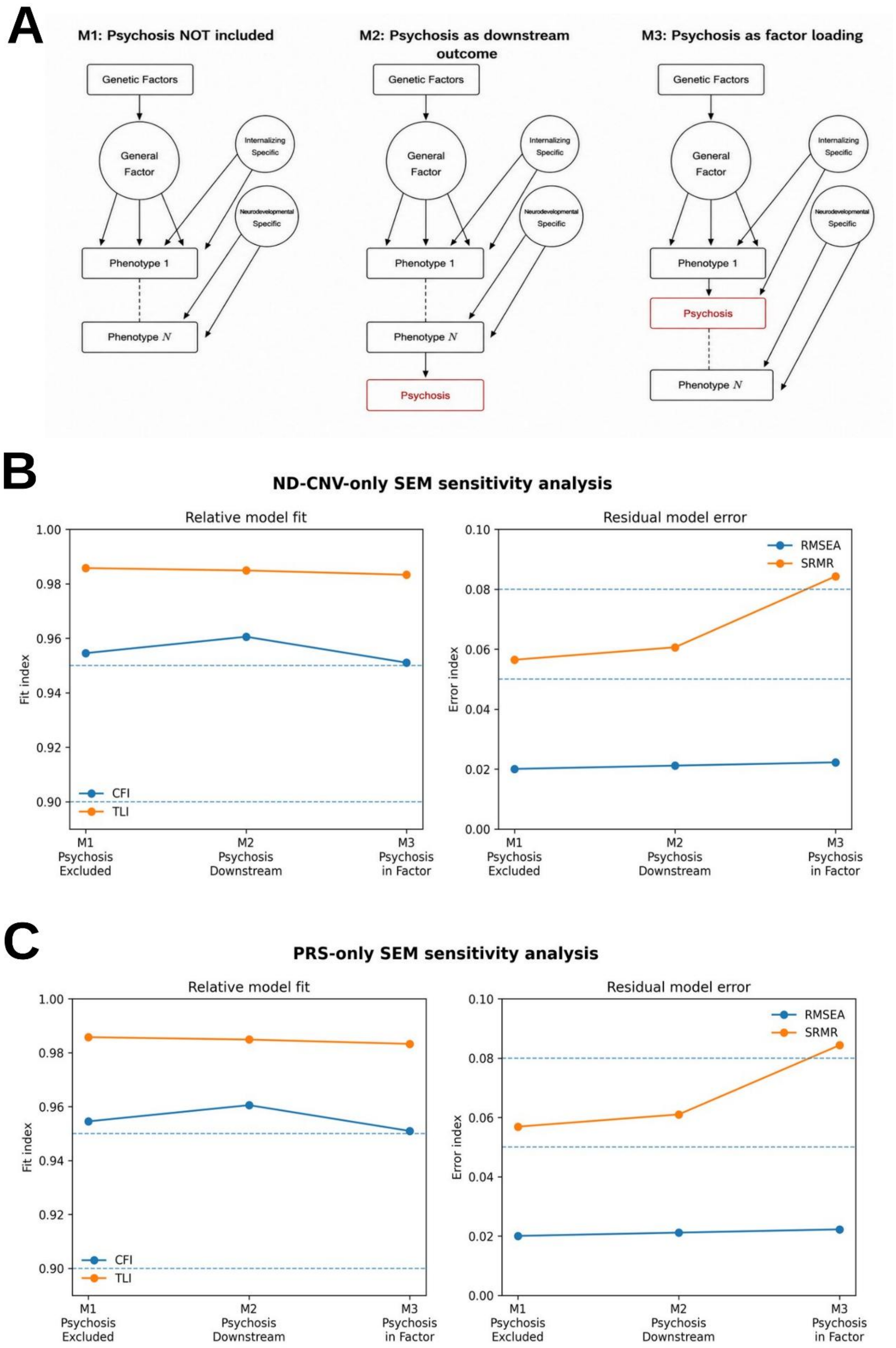
A
M1: Psychosis NOT included
M2: Psychosis as downstream outcome
M3: Psychosis as factor loading
Genetic Factors
General Factor
Internalizing Specific
Neurodevelopmental Specific
Phenotype 1
Phenotype N
Psychosis
B
ND-CNV-only SEM sensitivity analysis
Relative model fit
Residual model error
Fit index
Error index
CFI
TLI
RMSEA
SRMR
M1 Psychosis Excluded
M2 Psychosis Downstream
M3 Psychosis in Factor
C
PRS-only SEM sensitivity analysis
Relative model fit
Residual model error
Fit index
Error index
CFI
TLI
RMSEA
SRMR
M1 Psychosis Excluded
M2 Psychosis Downstream
M3 Psychosis in Factor

**Figure S7. Sensitivity of covariance-model fit to genetic predictor specification. (A)** Conceptual schematic of the three bifactor structural equation modeling (SEM) specifications evaluated in the primary analysis, corresponding to psychosis modeled as excluded from the general covariance structure (M1), downstream of general neuropsychiatric covariance (M2), or incorporated within the general covariance factor (M3), as also presented in Figure 3. **(B)** Model-fit sensitivity analysis using ND-CNV exposure alone as the genetic predictor. Relative fit indices (CFI, TLI; left) and residual error metrics (RMSEA, SRMR; right) are shown across the three SEM specifications. Dashed reference lines indicate commonly used thresholds for acceptable (0.90) and excellent (0.95) relative fit, and for close (0.05) and adequate (0.08) residual fit. Model-fit statistics (CFI, TLI, RMSEA, and SRMR) are shown for each SEM specification. **(C)** Model-fit sensitivity analysis using schizophrenia polygenic risk score (PRS) alone as the genetic predictor. Relative fit indices (CFI, TLI; left) and residual error metrics (RMSEA, SRMR; right) are shown across the same SEM specifications and threshold criteria. The overall pattern closely mirrored that observed in the ND-CNV-only analysis, with M2 again showing the highest relative fit among tested specifications.

### VI. External Supplemental Tables

**Supplementary Table 1: Modeled phenotype counts and prevalence.** Case counts, nonmissing sample sizes, and prevalence estimates for psychosis-spectrum outcomes and RDoC-informed neurobehavioral domains included in the primary analyses.

**Supplementary Table 2: Recurrent ND-CNV carrier prevalence across cohort strata.** Counts and prevalence of pooled, deletion-overlapping, and duplication-overlapping recurrent ND-CNV carriers stratified by predicted ancestry, age group, sex variable, and healthcare-utilization decile.

**Supplementary Table 3: Genomic annotation resources and participant-level overlap.** Definitions and sizes of the schizophrenia and neurodevelopmental gene sets used for annotation, together with the number and proportion of all participants and pooled, deletion-overlapping, and duplication-overlapping ND-CNV carriers whose intervals overlapped each resource.

**Supplementary Table 4: Phenotype-level associations of recurrent ND-CNV carrier status and schizophrenia PRS.** Complete primary and direction-specific logistic regression results across psychiatric and neurodevelopmental outcomes, including recurrent ND-CNV, schizophrenia PRS, interaction, and covariate estimates with odds ratios, confidence intervals, nominal p-values, and FDR-adjusted q-values.

**Supplementary Table 5: RDoC-informed dimensional association results.** Complete regression results for pooled and direction-specific recurrent ND-CNV, schizophrenia PRS, and interaction models across the six RDoC-informed neurobehavioral domains.

**Supplementary Table 6: Phenotype-level regression sensitivity analyses.** Results from alternative utilization adjustments, stricter repeated-diagnosis definitions, internalizing-adjusted models, and other prespecified perturbations used to evaluate the robustness of the primary phenotype-level associations.

**Supplementary Table 7: Schizophrenia polygenic risk score distributions.** Summary statistics for the standardized, residualized, and within-ancestry standardized schizophrenia PRS variables used in the regression and covariance analyses.

**Supplementary Table 8: CFA and SEM model-fit statistics.** Convergence and fit indices for the one-factor, correlated two-factor, and bifactor CFA models; primary psychosis-embedding SEM models; predictor-specific PRS-only and ND-CNV-only models; and reduced-factor sensitivity models.

**Supplementary Table 9: Key SEM path estimates and sensitivity analyses.** Unstandardized and standardized estimates for the principal schizophrenia PRS, recurrent ND-CNV, interaction, latent-factor, and psychosis paths across the primary SEM specifications and predictor-specific, reduced-factor, utilization, and leave-one-indicator-out sensitivity analyses.

Additional supporting materials, including complete machine-readable result tables, figure source data, analysis notebooks, code, quality-control outputs, and extended model results, are available through the accompanying Open Science Framework repository:

https://osf.io/4c58y/overview?view_only=4b449a32ac17473d89471a32f3c6c904.

Only aggregate, de-identified, and disclosure-compliant outputs are included.
